# Impact of Gamification in Behaviour Change Intervention: A Randomised Controlled Trial with YuLife’s Health and Wellbeing App

**DOI:** 10.64898/2026.05.31.26354543

**Authors:** Abbas Salami, Tasos Papastylianou, Osama Mahmoud, John Ronayne, Muhiba Rahimova, Bernard Fromson, Matt Doltis, Honor Bixby, Robert S. Stawski, Mariachiara Di Cesare

## Abstract

**Background:** Companies in the Health and Life Insurance space are increasingly turning to digital tools to promote healthier behaviours among their user base and reduce future health risks. This approach shifts insurers’ role from passive underwriters to partners in health management. These tools, often smartphone or wearable-tracker-based, enable real-time monitoring of behaviours (such as physical activity or meditation), providing fruitful targets for behavioural change interventions. Gamification, a Behavioural Change Technique with rich theoretical backing, is increasingly used in this context; however, despite its theoretical promise, current evidence remains mixed, and makes it hard to disambiguate its effect compared to more isolated financial incentives, the extent to which initial effects may be sustained over time, and how such changes in behaviour potentially translate to downstream health risk reductions.

**Objective:** This 9-month parallel-group, open-label Randomised Controlled Trial was designed to assess the causal impact of gamification in promoting health behaviours, independent of financial incentivisation. This was conducted in a real-world workplace setting, involving a cohort of participants using the YuLife Health and Wellbeing app, provided within an employer-sponsored group cover setting.

**Methods:** For the purposes of the RCT, the app was adapted such that gamification features could be turned on or off in a controlled manner, and in-app rewards in the form of “YuCoin” were adjusted between treatment groups to account for the effect of financial incentives. Following a baseline phase involving acquisition of baseline step estimates and questionnaire data, 1,288 participants – recruited from a number of companies partnered with YuLife, spanning various sectors – were randomised to gamified versus non-gamified versions of the app using stratified block-randomisation, and evaluated at specific milestones over a 9-month period, to enable comparison of short-term to long-term outcomes. The primary outcomes assessed were absolute differences in mean daily step count and engagement with the YuLife app. The data were analysed using Linear Mixed-Effects Models (LMMs). Additionally, a Cox Proportional Hazards model fitted to UK Biobank data was used to map step differences directly onto downstream health risks, and reductions were evaluated using an LMM. Further secondary outcomes (such as smoking and alcohol consumption) were also evaluated using non-parametric statistics.

**Results:** Compared with control, the gamified intervention was associated with greater mean daily steps throughout the study, with month × intervention interaction effects reaching one-sided 5% significance at months 3 (β=473.84, p=0.027), 5 (β=626.54, p=0.006), and 9 (β=480.91, p=0.033). Additionally, strong seasonal effects were identified, with fewer steps in Autumn (β≈−943.50, p<0.001) and Winter (β≈−1,145.45, p<0.001) versus Summer; higher baseline activity was a strong predictor of later activity (β≈0.85, p<0.001) and higher BMI was negatively associated with steps (β≈−60.84 per unit, p<0.001). For app engagement, month × intervention interactions were positive and significant from Month 3 onwards (Month 3 β=0.205, Month 5 β=0.182, Month 7 β=0.170, Month 9 β=0.175, all p<0.001), effectively showing sustained engagement while main milestone terms indicated declines in the control arm. Sensitivity analyses demonstrated the potential for baseline step inflation due to novelty effects, motivating repeating the step count analyses under an alternative baseline definition; this showed similar results, but with interaction effects achieving one-sided significance over all study milestones. Predicted partial-hazard analyses showed progressively larger month × intervention reductions in hazard, reaching one-sided significance at months 5 (coef=−0.018, p=0.016) and 9 (coef=−0.026, p=0.002). No significant intervention effects were observed for other secondary outcomes (e.g. smoking, alcohol) following Bonferroni-Holm correction.

**Conclusions:** Gamification elements can be an effective component in the context of digital interventions aiming to promote positive health behaviours, leading to improved engagement with the intervention and positive behavioural outcomes. Through progressive risk-reduction, even small but sustained improvements can be shown to meaningfully improve long-term health outcomes. Gamification is likely to add value to workplace health promotion initiatives, particularly for targeted short- to medium-term behavioural change interventions operating within a larger risk-management framework.

**Trial Pre-registration:** https://osf.io/926pd

## Introduction

Physical inactivity and sedentary lifestyles are among the most significant modifiable risk factors for all-cause mortality [1], contributing to around one in six deaths in the UK and increasing the risk for conditions such as cardiovascular disease, type 2 diabetes, musculoskeletal diseases, and obesity [2,3]. Physical inactivity has a substantial economic burden; the NHS spends almost £1 billion annually on inactivity-related ill health, while the broader cost to the UK economy, including loss of productivity, exceeds £7 billion a year [4]. In 2021, 36% of adults did not meet aerobic activity guidelines, with higher levels of non-adherence among women (41%) than men (30%), and significantly higher among the most deprived communities (47%) compared to the least deprived (32%) [5]. Mental ill-health, including stress, depression, and anxiety among working-age adults, presents a parallel concern. In 2023/24, these conditions affected almost 800,000 workers in the UK, leading to a loss of 16.4 million working days [6]. This translates into considerable costs to employers, estimated at around £51 billion annually, from absenteeism, reduced productivity (presenteeism), and staff turnover [7]. These figures underscore the urgent need for effective interventions targeting both physical and mental health within working populations.

Given this context, the workplace has emerged as a strategic setting for addressing lifestyle-related health risks [8,9]. In the UK, 87% of organisations offer some form of employee wellbeing program [10]. Often integrated into employer-sponsored insurance schemes, these programs aim to promote preventive health behaviours through tools such as health risk assessments, smoking cessation support, and gym discounts. Digital health technologies, particularly smartphone apps and wearable trackers, are increasingly central to these efforts [11,12]. These tools enable real-time monitoring of behaviours, such as physical activity or meditation, and enable scalable, personalised interventions [13]. A growing trend within this domain is gamification, or the integration of game elements into non-game contexts [14]. Based on psychology and behavioural science theories, gamified health programs often include step challenges, reward points, virtual badges, or social competitions, aiming to increase user motivation by making healthy behaviours more engaging, enjoyable, and rewarding [15]. In contrast to traditional approaches relying on long-term rational appeal, gamified interventions provide immediate gratification and a sense of progress, which may help overcome procrastination and other behavioural barriers [12]. These programs can yield meaningful benefits for both employees and employers through improved health, reduced absenteeism, and potentially favourable returns on investment [16].

Despite its theoretical promise, the evidence base for gamified health interventions remains mixed [14,17–19]. Several studies have shown short-term behavioural improvements associated with gamification [20,21], suggesting that gamification may be effective in boosting short-term physical activity across a range of populations, including both healthy individuals and those managing chronic conditions [22].

However, many studies combine gamification with monetary incentives, making it difficult to isolate the contribution of game elements alone [23,24]. In addition, there is a lack of evidence on the long-term impact of gamification on health behaviours, with most trials spanning 8 to 12 weeks, making it difficult to assess whether behaviour changes are sustained over time [18]. Moreover, there is limited evidence on how improvements in health behaviours, such as increased physical activity, translate into meaningful reductions in health risks [21,25]. Employers and insurers implementing gamified wellness programs aim to reduce health risks, lower healthcare claims, and improve workforce productivity, but few rigorous evaluations have directly examined these outcomes. Even large, high-quality RCTs have shown mixed results in the broader wellness literature. For instance, an 18-month cluster RCT of a comprehensive wellness program in a large U.S. company found no significant effects on health measures, healthcare costs, or absenteeism, despite reported improvements in health behaviours [26].

In light of these gaps, this study evaluates the isolated impact of gamification on employee health behaviour and health outcomes in a real-world workplace setting. We conducted a large-scale, 9-month Randomised Control Trial (RCT) involving over 1,000 UK employees using the YuLife mobile health and wellbeing app. This study provides novel evidence on the standalone impact of gamification over an extended period in a UK workplace context. To our knowledge, this is the first large-scale RCT to isolate the effects of gamification from those of financial incentives while evaluating behavioural and risk-related outcomes. The findings aim to inform future employers, policymakers, and insurers’ decisions about whether gamified health apps can meaningfully contribute to employee wellbeing and, more generally, to population health.

## Methods

### Study Design

YuLife Ltd is a UK-based “insurtech” [27] company that provides life insurance and employee wellbeing benefits through a technology-driven platform. The heart of this platform is the YuLife Health and Wellbeing app (or simply, the “YuLife app”), which encourages users to engage in healthy behaviours, such as walking, cycling, and meditation, through a gamified interface featuring virtual points, levels, avatars, streaks, and team challenges. We conducted a parallel-group [28], open-label [29] RCT to assess the causal impact of implementing gamification in a health behaviour change intervention delivered through the YuLife mobile app on employee health behaviour and wellbeing outcomes, independent of financial incentivisation. To isolate the effects of gamification features (such as levels, challenges, and leaderboards) on behavioural and mental health outcomes, two versions of the YuLife mobile app were used: a fully gamified version (intervention group) and a version stripped of all game elements (control group), while maintaining similar monetary incentive exposure in both arms. The trial period lasted 9 months, during which participants used their assigned app version with no planned follow-up beyond the intervention window. Participants were recruited from several companies partnered with YuLife. Due to staggered company onboarding, randomisation was conducted within each company to ensure balanced representation across intervention and control arms. While randomisation was performed at the individual level, stratification was applied based on age, gender, and baseline physical activity (daily step count) to control for known confounding factors. As this was a real-world deployment, blinding was not feasible. Participants could clearly distinguish between the two versions of the YuLife app based on their interfaces and features. Additionally, although the intervention group earned extra YuCoin (the app’s internal reward currency) through engagement with gamified features, the control group received an equivalent amount of YuCoin each week on average, ensuring equal exposure to monetary incentives. This parity in incentives was designed to isolate the incremental effect of gamification. Both groups received the same level of user support and app functionality outside of gamification, and the intervention was delivered entirely through the YuLife app. This study was registered with the Open Science Framework (OSF) on 16 April 2024 [30], before any participants were enrolled. The study protocol was reviewed and approved by the University of Essex Ethics Sub Committee 1 (ETH2324-0764). All data were collected and managed in accordance with ethical standards and data protection regulations.

### Participants

Participants were employees from six companies that were prospective clients of YuLife. The six companies operate across diverse sectors: information and communication; transport and storage; wholesale and retail trade; administration and support services; and finance. All employees at each participating company were invited to participate in the study. Invitations were distributed via the company’s internal communications channels and included a participant information sheet and an electronic consent form. Participation was voluntary and conditional upon providing informed consent via a secure digital platform (SurveyMonkey). Participants were eligible if: 1) were 18 years or older; 2) employed; 3) owned a smartphone compatible with the YuLife app; 4) capable to engage in basic physical activity (e.g., walking); 5) had no prior experience of using the YuLife app; 6) not enrolled in another health behaviour study; 7) had no pre-existing medical conditions that could be exacerbated by increased physical activity.

### Data Collection

Data were collected using a combination of automated app tracking, wearable device integration, and questionnaires administered both in-app and prior to the study. Data collection occurred in two phases: a baseline phase and a trial phase. During the 2-week baseline period, all participants used a non-gamified version of the app without weekly YuCoin rewards. This phase served two purposes: to allow participants to familiarise themselves with the app and to establish baseline measures of daily step count. Baseline step count was calculated as the mean daily steps over this period and was used for stratified randomisation and as a covariate in subsequent analyses. Participants were also invited to complete a baseline health questionnaire during this period via a secure SurveyMonkey link sent to their work email. Step counts were recorded automatically using smartphone sensors; where participants connected a wearable device, daily step counts were derived by selecting the higher value recorded between the smartphone and the wearable device to reduce potential underestimation due to incomplete smartphone carriage. App engagement was defined as the proportion of active days over a representative period of interest, namely the baseline period when evaluating baseline engagement, or a 30-day window when evaluated at follow-up points. An active day was defined as any calendar date on which the user had at least one record in the app. The outcome, engagement percentage, is calculated as the number of distinct active days divided by the total number of days in the window, expressed as a proportion between 0 and 1. During the trial phase, participants were invited to complete follow-up health questionnaires at 3, 6, and 9 months, administered via email with reminder notifications delivered through the app. The questionnaires captured a range of secondary outcomes, including self-reported sleep duration, body mass index (BMI), smoking and alcohol use, perceived stress, anxiety, depression, work productivity, and job satisfaction. Standardised instruments were used for mental health outcomes, including the Perceived Stress Scale (PSS-4) [31], Generalised Anxiety Disorder scale (GAD-2) [32], and Patient Health Questionnaire (PHQ-2) [33] (see Multimedia Appendix). In addition to these scheduled assessments, the app included optional in-app questionnaires, referred to as Dynamic Health Questionnaires (DHQ), which enabled the collection of additional self-reported health data while providing personalised wellbeing recommendations and small in-app rewards for completion (additional details and example screenshots are provided in the Multimedia Appendix).

### Randomisation

Participants were randomised to one of two groups, intervention (gamified app) or control (non-gamified app), using a stratified randomisation. The aim was to ensure balanced distribution across key confounding variables and company size. Randomisation was conducted at the individual level, with participants stratified by: age (≤33, 34–46, >46); gender (male, female); and baseline physical activity (≤5,000 steps/day, 5,001–9,000 steps/day, >9,000 steps/day). Within each participating company, randomisation batches of varying sizes were created to accommodate differences in company size and staggered entry. The batch sizes ranged from 80 to 300, depending on the number of eligible users in each company or cohort. Randomisation was implemented using a Python-based script, developed and executed by the academic research team at the University of Essex. The allocation sequence was generated and applied after the 2-week baseline phase, once participants had completed their setup and baseline step count data were available. Although the study was open-label and participants were not blinded to the version of the app they received, allocation concealment was preserved. The group assignment occurred automatically, without involvement from participants, company staff, or any individual who could influence enrollment. This approach minimised potential selection bias while preserving the integrity of the randomisation process.

### Intervention Overview

#### Intervention Group

Participants in the intervention group received the full version of the YuLife app, which incorporated a comprehensive set of gamified features designed to promote sustained engagement and physical activity. While all participants could passively earn the app’s virtual currency (YuCoin) through health-related activities, including daily step count, meditation duration, and cycling distance, the intervention group had access to additional reward opportunities embedded within gamified mechanisms. These mechanisms included levels, challenges, duels, leaderboards, badges, streaks, and avatar-based progression, all designed to reinforce engagement in positive health behaviours. Challenges were implemented as goal-oriented tasks (e.g., completing a predefined number of steps within a specified period, completing workout sessions, or meditating for a target duration). The gamified experience was further structured through themed “universes”, providing a narrative context and a sense of progression within the app.

In addition, participants had access to *Yudoku* (YuLife’s version of Sudoku), a daily brain-training activity offering additional rewards, limited to one completion per day. Periodic in-app events were also introduced, consisting of time-limited challenges (e.g., achieving specific activity targets within a defined timeframe) with additional incentives.

Alongside these elements, participants retained access to core functionalities available to both study groups, including virtual general practitioner (GP) support, an Employee Assistance Programme (EAP), wellbeing product discounts, the DHQ, and a mood monitoring feature. A detailed description of the gamification components (including how these map onto the Behavioural Change Technique taxonomy [12,34]), along with representative screenshots of the app interface, is provided in the Multimedia Appendix.

#### Control Group

Participants allocated to the control group received a version of the YuLife app with an identical user interface and overall structure, but with all gamified features disabled. Similar to the intervention group, participants could earn YuCoin passively through engagement in health-related activities, including daily step count, meditation duration, and cycling distance.

To ensure comparability in overall reward exposure between groups, participants in the control group received an additional weekly allocation of YuCoin equivalent to the average additional amount earned by intervention participants through interaction with gamified elements.

The control group retained access to the same core, non-gamified services as the intervention group, including passive activity tracking, virtual GP support, an EAP, wellbeing product discounts, the DHQ, and mood monitoring.

### Study outcomes

This trial was designed to evaluate the impact of gamification on physical activity and app engagement, alongside a set of exploratory behavioural, mental health, and work-related outcomes.

#### Primary Outcomes

The primary focus of the trial was to assess the effect of gamification on two key outcomes:

1. *Physical Activity* We sought to assess whether participants using the gamified version of the YuLife app will demonstrate a greater absolute increase in average daily step counts from baseline, both in the short term and sustained over time, compared to those using the non-gamified version of the app. The relevant null and alternative hypotheses are:

a. Null Hypothesis (H0): There is no difference in step counts between the control and intervention groups, when evaluated at the 3-month milestone. Alternative Hypothesis (H1): The intervention group will have a higher mean daily step count compared to the control group, when evaluated at the 3-month milestone (one-sided alpha = 0.05).
b. Null Hypothesis (H0): There is no difference in step counts between the control and intervention groups, when evaluated at the 9-month milestone. Alternative hypothesis (H1): The intervention group will have a higher mean daily step count compared to the control group, when evaluated at the 9-month milestone (one-sided alpha = 0.05).
2. *App Engagement* We sought to assess whether participants in the gamified group would exhibit higher app engagement across the duration of the intervention, relative to the control group. The relevant null and alternative hypotheses are:

a. Null Hypothesis (H0): There is no difference in engagement between the control and intervention groups, when evaluated at the 3-month milestone. Alternative Hypothesis (H1): The intervention group will have a higher engagement compared to the control group, when evaluated at the 3-month milestone (one-sided alpha = 0.05).
b. Null Hypothesis (H0): There is no difference in engagement between the control and intervention groups, when evaluated at the 9-month milestone. Alternative Hypothesis (H1): The intervention group will have a higher engagement compared to the control group, when evaluated at the 9-month milestone (one-sided alpha = 0.05).

#### Secondary Outcomes

Secondary outcomes included a range of self-reported behavioural, mental health, and work-related measures collected via periodic questionnaires administered at three-month intervals. These included sleep quality, BMI, smoking and alcohol consumption behaviours, perceived stress, anxiety, depression, and work-related outcomes (e.g. productivity and job satisfaction).

Given the lower response rates for questionnaire-based measures, analyses of secondary outcomes were considered exploratory and are reported in the Multimedia Appendix where applicable.

### Statistical Analysis

#### Power calculations

A priori power analysis was conducted using G*Power 3.1 [35] to determine the minimum required sample size for detecting a meaningful effect on the primary outcome, the mean daily step count. The analysis was based on a two-group comparison of independent means, assuming a Cohen’s *d* of 0.2, corresponding to an approximate 10% increase in average daily step count following the use of the YuLife app. The power level was set at 0.80, with a one-sided alpha level of 0.05 and an allocation ratio of 1:1 between intervention and control groups. Under these assumptions, the required sample size was calculated to be approximately 310 participants per group or 620 in total. To account for an anticipated dropout rate of roughly 40%, the recruitment target was increased to at least 1,000 participants.

#### Per-protocol criteria and final sample

Analyses were conducted using a per-protocol approach. Participants were included only if they adhered to the study protocol and provided valid data. This required downloading and using the app, as it was both the intervention platform and the primary source of outcome data. Participants also had to meet predefined baseline criteria, including timely synchronisation of wearable data prior to randomisation to ensure that step counts reflected actual behaviour during the baseline period. Individuals were excluded if they did not use the app, failed to synchronise within the required window, or had insufficient baseline or follow-up data. These criteria reduced bias from irregular synchronisation and ensured reliable baseline measurement. The final sample, therefore, represents participants who engaged with the intervention and maintained adequate data coverage, allowing estimation of the intervention effect under conditions of adherence rather than simple assignment.

#### Data Handling

Missing data points were handled according to the nature of each variable. For step count, if no data were recorded in the 30-day window preceding each checkpoint (3, 5, 7, and 9 months), the corresponding mean daily step count for that time point was considered missing. However, periods of inactivity (e.g., no app engagement) were not treated as missing data but rather as valid indicators of zero engagement.

Outlier detection for step count data was conducted at the individual level. Any daily step count below 100 steps or above 50,000 steps was considered implausible and excluded from the dataset. If such outliers occurred within a broader sequence of valid step records for that participant, the missing values were later interpolated using the matrix completion approach described below.

A two-step imputation strategy was employed to address missing step data. First, matrix completion by iterative soft thresholding of singular value decomposition (SVD) from fancyimpute package [36] was used to interpolate missing individual step count days without extrapolating beyond the participant’s recorded range. Next, Multivariate Imputation by Chained Equations (MICE) [37] was applied to estimate the remaining missing mean daily step counts at different checkpoints. The imputation model included demographics (age, gender, ethnicity, weight, height, BMI), step counts and engagement proportions at the various milestones, and a baseline season factor.

For secondary outcomes derived from health questionnaires, missing values were recorded for items not completed by participants. Where possible, responses from the DHQ were used to supplement or impute missing values for selected secondary outcomes, as they contained overlapping items with the full health questionnaires and demonstrated higher participant completion rates.

#### Primary outcome analyses

To assess the impact of gamification on physical activity and engagement, we fitted linear mixed-effects models (LMMs).

The LMM model for mean daily steps included fixed effects for time treated as a categorical variable (baseline, 1 month, 3 months, 5 months, 7 months, and 9 months), treatment group (intervention vs control), season (to account for seasonality and staggered entry), baseline-centred steps, BMI, and the time × group interaction to evaluate differential changes over time between the two groups. Random intercepts were included for participants nested within their respective companies to account for repeated measurements and clustering within organisations. Reference categories were time = 0 (i.e., the study baseline), Control group, and Summer season. Baseline-centred steps were defined as each participant’s baseline steps minus the cohort mean, allowing post-baseline group differences to be estimated after accounting for any pre-existing differences. The model also adjusted for gender and age group.

The LMM model for app engagement included fixed effects for time (baseline, 3 months, 5 months, 7 months, 9 months), treatment group, season, baseline-centred engagement, and the time × group interaction, with random intercepts for participants nested within companies. Baseline-centred engagement was defined as each participant’s baseline engagement minus the cohort mean, as with steps. Reference categories were time = 0, Control group, and Summer season.

In both cases, for each fixed effect other than time × intervention interaction effects, we report coefficient estimates along with their associated two-sided p-values; for time × intervention interaction effects we report one-sided p-values instead, as per our null and alternative hypotheses. Data cleaning and preprocessing were performed using Python (version 3.12). All statistical analyses were performed using R (version 4.4.2), utilising the lme4 [38], lmerTest [39], ordinal [40], and mice packages.

Additional visualisations included raw (unadjusted) and model-adjusted outcomes per milestone per treatment group, and the seasonal pattern in raw step counts.

#### Sensitivity analyses

To account for the potential impact of strict per-protocol criteria retaining only a very restricted sample out of the total randomised participants, we conducted sensitivity analyses to characterise potentially excluded participants and evaluate how the intervention effect might be impacted if we could account for these individuals.

For the purposes of the study, participants were intended to download the app and synchronise their devices at the start of the baseline period, as downloading the app alone does not ensure device synchronisation or step data collection. This entails that, ideally, given the two-week baseline, the activation delay, defined as the number of days between first device synchronisation and the randomisation date, would be expected to cluster tightly around 14 days, as expected by the protocol and corresponding instructions given to participants. In the per-protocol analysis, only this tight cluster was considered for the analysis of primary outcomes. Here, we account for a potentially wider distribution of activation delays, reflecting the potential for larger deviations in synchronisation behaviour in the studied population, and outline a further analysis approach able to account for such a wider distribution, to work around the case of a highly restricted sample resulting from the strict per-protocol criteria.

As users cannot view their steps or earn rewards until they synchronise their device, the first step in this analysis involves defining which temporal anchor (T=0) serves best for measuring baseline activity. For this purpose, we consider three temporal anchor candidates:

- First App Open (defined as the date a user first logs in to the app); under this anchor, features remain locked, and no tracking data is available until sync.
- Split Date (defined as the date on which the participant was randomised into a treatment group); this reflects a system event, independent of user readiness
- Sync Date (defined as the first passive step recorded with app status as active); this reflects a valid behavioural start.

Since the choice of temporal anchor may also lead to potential contamination from novelty effects, we further consider a classification of each participant into one of four cohorts based on their syncing behaviour (and in particular, the number of valid data points before the Sync Date), as shown in Table 1.

**Table 1.** Participant classification criteria.

| **Cohort Name** | **Criterion** | **Rationale** |
| --- | --- | --- |
| De Novo Trackers | <10 days of tracking history before Sync Date | Insufficient pre-intervention data to establish baseline |
| Quitters | Last recorded activity <30 days after Sync Date | Early dropouts cannot contribute to longitudinal analysis |
| Fragmented | Any data gap >60 consecutive days post-sync | Valid baseline but inconsistent follow-up tracking |
| High-Quality Sync | ≥10 days pre-sync AND consistent post-sync engagement with no gaps >60 days | Gold standard: valid baseline + consistent engagement |

We therefore consider here on this basis, a revised definition of baseline step count, grounded in users’ physical activity prior to exposure to the app and its novelty effect. Specifically, the baseline is measured using a 30-day pre-sync window (with a minimum of 10 valid days), reflecting natural behaviour before users can view their steps or earn in-app rewards. This approach requires excluding De Novo Trackers from analyses requiring valid pre-intervention baselines, alongside Quitters and participants who never synchronised their devices.

Armed with this information, we then repeat the mean daily steps analysis as before, using the revised baseline definition and resulting cohort after retaining high-quality and fragmented users only.

#### Health Risk Modelling

To evaluate the longer-term clinical relevance of the behavioural changes observed in the trial, we estimated participants’ predicted all-cause mortality risk using a Cox proportional hazards model [41] trained on the UK Biobank. The UK Biobank is a large-scale, population-based biomedical database comprising over half a million UK residents, with comprehensive demographic, behavioural, and health outcome data [42]. Around 100,000 Biobank participants also wore wrist accelerometers for 7 days, providing objectively measured step data. Using this subset, we trained a Cox model to predict time to all-cause mortality based on demographic and behavioural predictors available in both the UK Biobank and our RCT, including age, sex, average daily step count, sleep duration and BMI. Time to event or censoring was calculated from the date of activity measurement to the end of follow-up in the Biobank. All model development was conducted using the lifelines package in Python [43] (see Multimedia Appendix for the full details of model development and validation). We then applied the Biobank-trained model to the RCT cohort, using each participant’s behavioural profile at baseline and follow-up checkpoints. A predicted individual hazard ratio was estimated at each time point for each participant. Finally, to quantify the intervention’s effect on predicted health risk, we reported the average hazard ratio for each time point in the intervention and control groups. To statistically evaluate the impact of the intervention on health risk reduction, we also used LMMs similar to previous analyses; the model, fitted via REML, included fixed effects for time, treatment group, age, and sex, together with the time × group interaction terms, and random intercepts for participants. Reference categories were time = baseline, Control group, and Female.

## Results

A total of 1,952 employees across six companies were invited through their company networks to participate in the study, with recruitment conducted on a staggered basis between May and July 2024. The invitation provided participants with information about the study and a consent form, as well as an invitation to complete the baseline health questionnaire and download the YuLife mobile app. 1,926 employees were considered for eligibility following consent (1,826 explicit via returned consent forms, and 100 implicit through continued engagement with the app following study onboarding but with no consent form returned) as per the approved ethics guidelines. Of these participants, 246 individuals were excluded because they reported injuries, musculoskeletal conditions, or other health issues that could limit their physical activity; a further 382 participants who did not download the app were also excluded from the trial, as app usage was essential for both delivering the intervention and collecting outcome data. Finally, a further 10 participants were excluded from treatment allocation because their first login occurred outside the time window that would allow them to complete within the study’s allocated timeframe. This resulted in a final sample of 1,288 individuals, randomly assigned to the Control (660) and Intervention (628) treatments.

Following randomisation, we applied strict per-protocol criteria requiring device synchronisation within a ±24-hour window of the 14-day pre-randomisation checkpoint (i.e., 13–15 days before the split date). This timing requirement ensures that baseline step measurements reflect genuine behavioural engagement during the study’s baseline phase rather than backfilled historical data retrieved from device memory upon initial synchronisation. Of the 1,288 randomised participants, 859 were excluded from the per-protocol analysis: 322 late syncers (synchronised fewer than 13 days before randomisation, providing insufficient real baseline data), 256 early syncers (synchronised more than 15 days before randomisation – which risks introducing potential novelty effect contamination into baseline measurements; see later sections), 149 post-split syncers (synchronised after randomisation, precluding any pre-intervention baseline), 92 participants who never synchronised their device, and 39 participants with insufficient baseline data (fewer than 10 days of step recordings within the baseline window). This resulted in a final per-protocol sample of 427 participants (203 Control, 224 Intervention) for the primary analysis (Figure 1).

**Figure 1.**
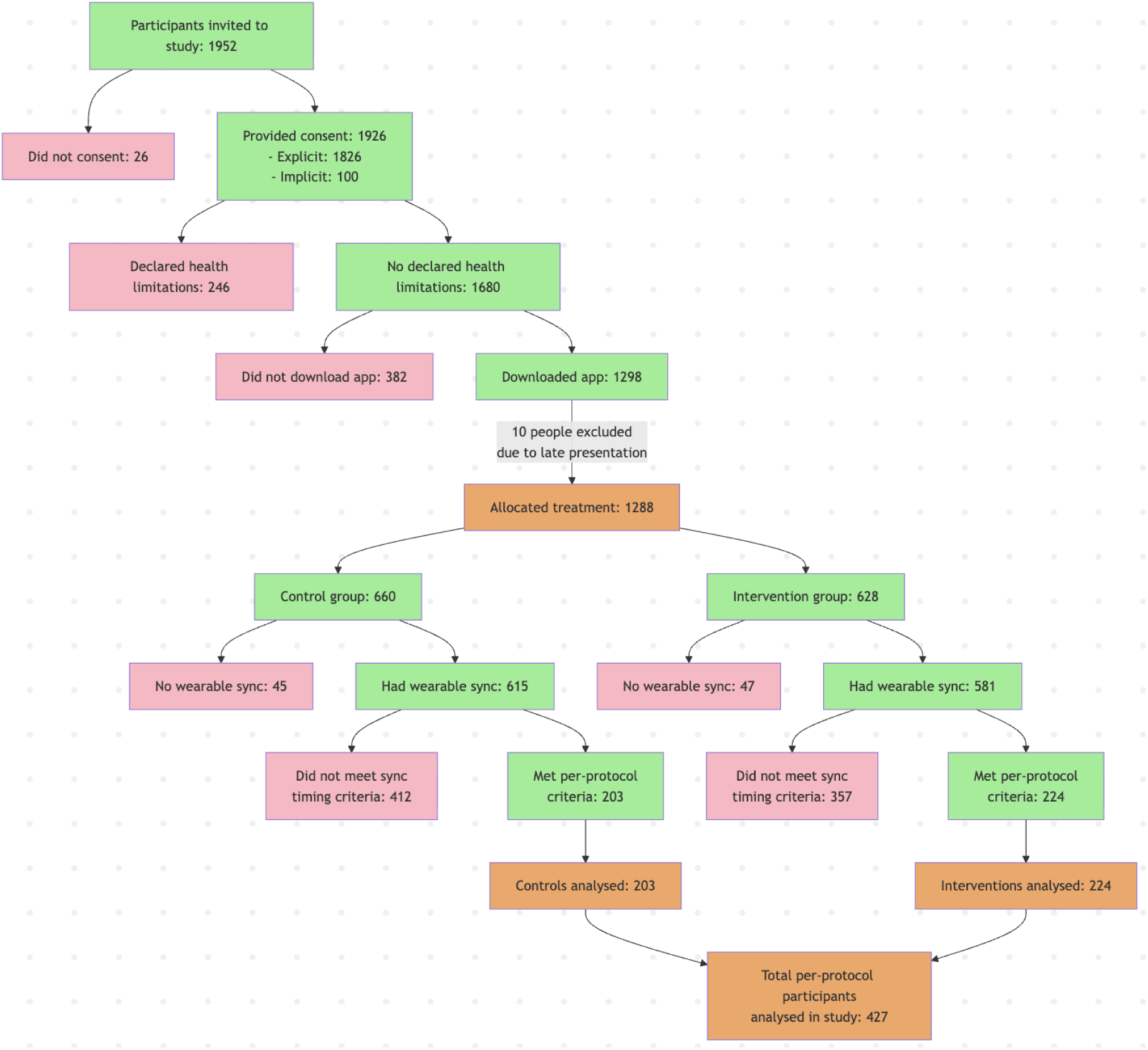
Participant flow diagram. Green boxes indicate included participants at each stage; pink boxes indicate exclusions; orange boxes indicate key milestones (allocation, final analysis sample). Per-protocol criteria required wearable synchronisation within ±24 hours of the 14-day pre-randomisation checkpoint to ensure baseline measurements reflect genuine behavioural engagement rather than backfilled historical data.

Figure 2 presents participant retention rates throughout the study. Of the 427 per-protocol participants, 291 (68.1%) maintained step data recording through month 9, with similar retention rates between Control (143/203, 70.4%) and Intervention (148/224, 66.1%) groups. Health questionnaire response rates showed expected attrition: 390 participants (91.3%) completed the baseline questionnaire, reduced to 200 (46.8%) at 3 months, 168 (39.3%) at 6 months, and 185 (43.3%) at 9 months. Response rates were balanced between groups throughout the study.

**Figure 2.**
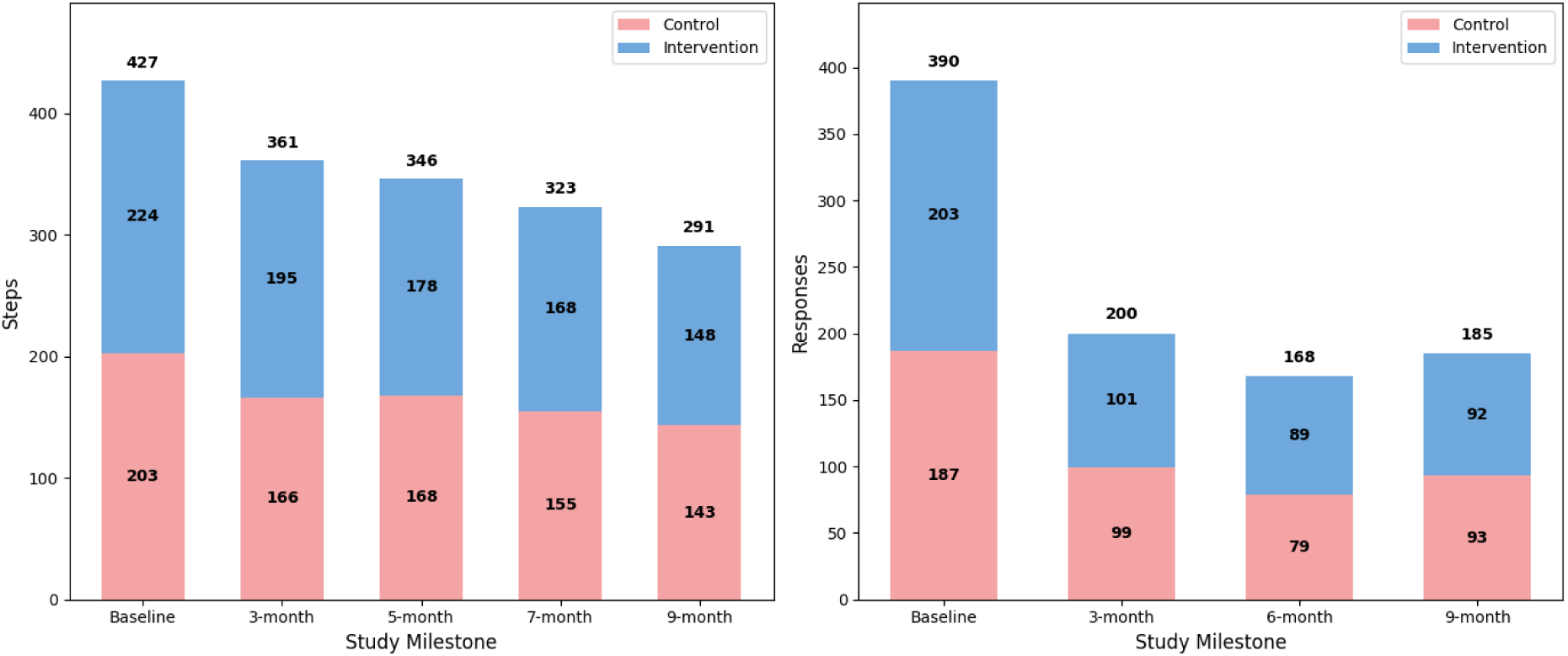
Retention of participants: step activity (left panel) and response rates to the health questionnaire (right panel).

### Baseline Characteristics

Baseline demographic and behavioural characteristics (age, gender, and average daily steps) are presented in Table 2. No statistically significant differences were identified between the intervention and control group (p>0.05), indicating that the randomisation process achieved adequate balance across the key variables.

**Table 2.** Baseline characteristics by group.

| Variable | Intervention (n=224) | Control (n=203) | Total (n=427) | p-value* |
| --- | --- | --- | --- | --- |
| Age (mean, SD) | 40.9 (11.4) | 39.9 (11.3) | 40.4 (11.4) | 0.389 |
| Gender (% female) | 105 (47%) | 107 (53%) | 212 (50%) | 0.406 |
| Average steps (mean, SD) | 7881.2 (3347) | 7883.8 (3738) | 7882.4 (3534) | 0.994 |

\* p-values from independent samples t-test (age, baseline steps) or chi-squared test (gender).

### Primary Outcomes (Per-Protocol Analysis)

#### Mean Daily Steps

Trends in the unadjusted average daily step counts for both treatment groups over the study period are shown in Figure 3A. Visual inspection indicates that the intervention group maintained higher step counts than the control group for most of the trial. Both groups also exhibit a noticeable decline in steps during the autumn and winter months (approximately months 5–7) suggesting a clear seasonal pattern (see Figure 3.1 in the Multimedia Appendix).

**Figure 3.**
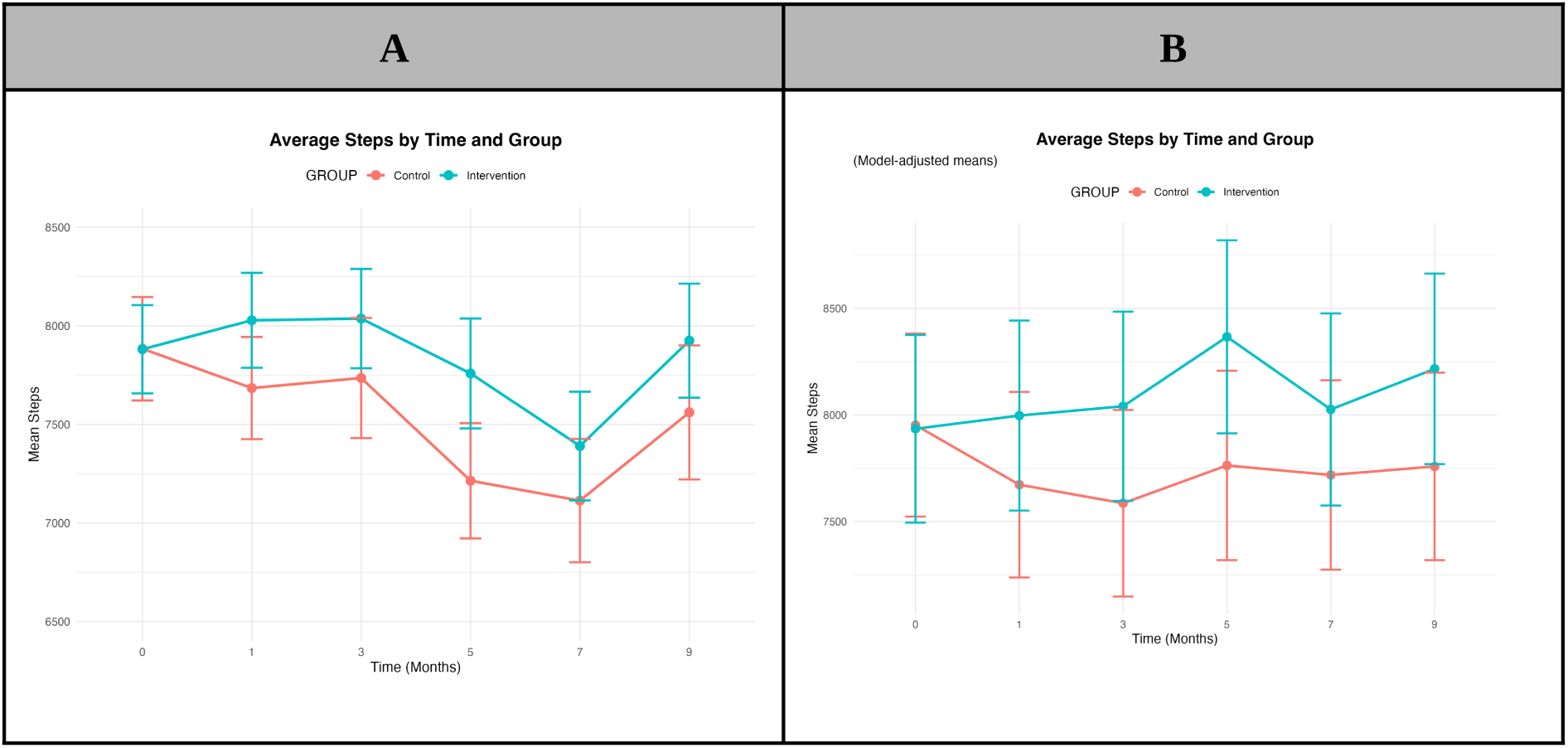
(A) Unadjusted and (B) model-adjusted average daily step counts by treatment group across study milestones (adjusted for seasonality). Error bars represent 95% confidence intervals.

The results showed that the two treatment groups did not differ meaningfully at baseline after adjustment (Intervention group β = −31.78, p = 0.872), indicating balance between groups. In the Control group, mean daily steps were generally lower than baseline at each milestone, though none of these differences reached statistical significance (month 1: β = −280.88, p = 0.135; month 3: β = −368.66, p = 0.060; month 5: β = −194.99, p = 0.388), month 7: β = −237.37, p = 0.285; month 9: β = −197.57, p = 0.335).

Conversely, the time × Intervention interaction terms were positive at every follow-up (and generally, with more positive values than the respective negative values in the control group for the same milestones), indicating higher step counts in the Intervention group relative to Control (month 1: β = 341.98, one-sided p = 0.071; month 3: β = 473.84, one-sided p = 0.027; month 5: β = 626.54, one-sided p = 0.006; month 7: β = 330.64, one-sided p = 0.095, month 9: β = 480.91, one-sided p = 0.033), with months 3, 5, and 9 reaching statistical significance.

Baseline-centred steps was a strong predictor of subsequent steps (β = 0.8531, p < 0.001), indicating that participants with higher baseline activity tended to record more steps at follow-up. Seasonal effects were also evident: Autumn (β = −943.50, p < 0.001) and Winter (β = −1,145.45, p < 0.001) were associated with substantially fewer mean daily steps compared with Summer; Spring showed a non-significant reduction (β = −194.76, p = 0.168). BMI was associated with lower step counts (β = −60.84 per unit increase in BMI, p < 0.001).

In summary, the gamified intervention was associated with higher mean daily steps relative to control at all follow-up milestones, with the strongest evidence at month 5, but otherwise showing evidence of a sustained effect throughout the study. Baseline activity, season, and BMI were also important predictors of subsequent step counts. The full model results are provided in the Multimedia Appendix (see Table 2.1). We note that the equivalent analysis using imputation of missing data (as discussed in the Data Handling section, reflecting a modified intention-to-treat analysis), yielded similar results for the time × intervention interaction effects (but not necessarily for the controls, which showed both negative and slightly positive effects, rather than purely negative); these are provided in Box 4 of the Multimedia Appendix.

Figure 3B presents model-adjusted average steps (estimated marginal means) for both treatment groups over the study milestones.

#### App Engagement

The results showed that the two treatment groups did not differ significantly at baseline after accounting for pre-existing engagement differences (Intervention group β = 0.018, p = 0.531), indicating adequate balance between groups. In the Control group, engagement declined substantially after randomisation, with reductions of approximately 20–26 percentage points from month 3 onwards (month 3: β = −0.218, p < 0.001; month 9: β = −0.255, p < 0.001), corresponding to 6-8 days in the 30-day analysis window. This suggests that, in the absence of gamification, app engagement tended to decrease markedly over time.

The time × Intervention interaction terms were positive and statistically significant at every follow-up point. The estimated treatment effects were +20.5 percentage points at month 3, +18.2 percentage points at month 5, +17.0 percentage points at month 7, and +17.5 percentage points at month 9, all with one-sided p < 0.001. These effects were similar in magnitude, and opposite in direction, to the corresponding time effects observed in the Control group, suggesting that the intervention largely offset the post-randomisation decline in engagement. Importantly, the effect was sustained through to month 9, indicating that the intervention did not simply produce a short-term activation effect.

Baseline engagement was a strong predictor of subsequent engagement (β = 0.732, p < 0.001), highlighting substantial between-user differences in app usage patterns. Seasonality was also observed, with Spring associated with higher engagement compared with Summer (+7.5 percentage points, p < 0.001). Overall, these results provide evidence that the gamified version of the YuLife app significantly increased and sustained app engagement relative to the non-gamified control condition across the full 9-month study period. Figure 4 shows unadjusted and model-adjusted average engagement (estimated marginal means) for both treatment groups over the study milestones. The full model results are provided in the Multimedia Appendix (see Table 2.2).

**Figure 4.**
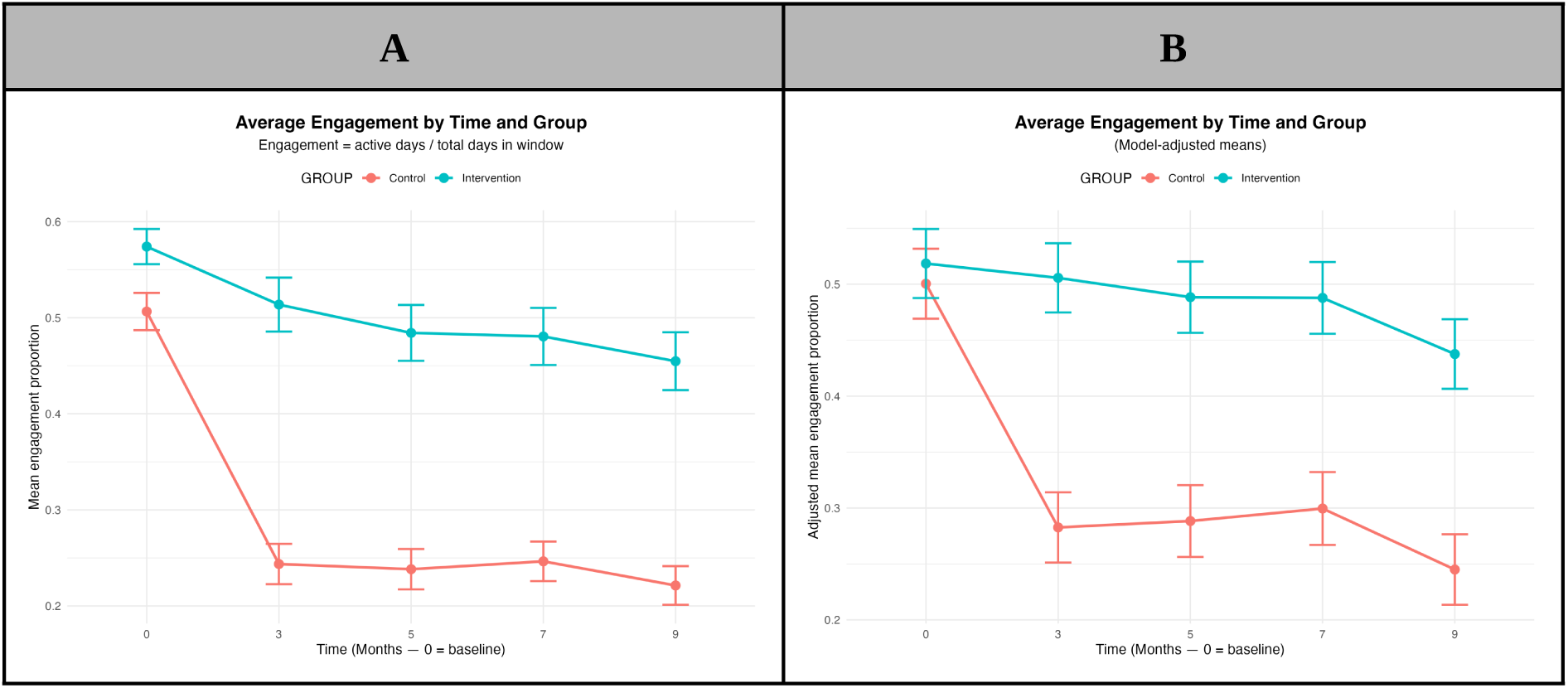
(A) Unadjusted and (B) model-adjusted (EMM) mean engagement by time × group (±1 SE), holding season and baseline at reference values.

### Sensitivity Analysis: Novelty Effect and Alternative Baseline

The strict per-protocol criteria retained only 33% of randomised participants (427 of 1,288). Examination of the complete randomised sample revealed substantial heterogeneity in the timing of participants’ synchronisation of their mobile or wearable devices (Figure 5).

**Figure 5.**
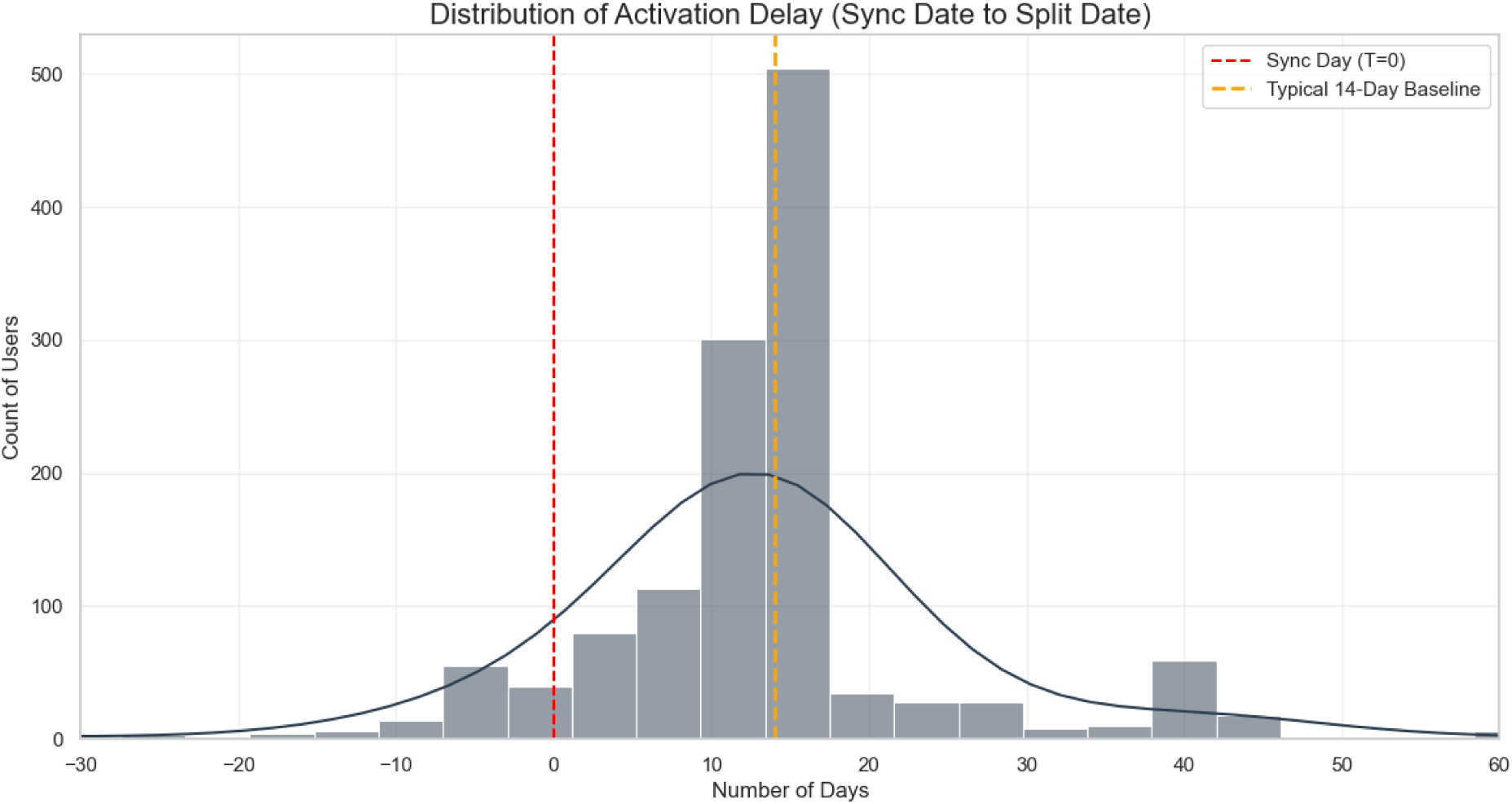
Distribution of activation delay (Sync Date to Split Date). The histogram shows when participants first synchronised their devices relative to their randomisation date. The red dashed line marks the sync day (T=0), while the orange dashed line indicates the typical 14-day baseline window specified in the study protocol. Note: A small number of negative values appears here due to the fact that participants were allocated treatment in batches to account for company logistics and staggered entry. In a small number of participants, stratification occurred on the basis of secondary data from the Baseline questionnaire, with those participants syncing at a later date, resulting in this negative “Sync-to-Split” difference.

Figure 5 shows that participants’ sync timing spans nearly 90 days, from 30 days before to 60 days after their intended baseline period. For the majority of participants who synced late or after randomisation, their apparent “baseline” data is either backfilled from device memory or simply does not exist as a genuine pre-intervention measurement.

#### Novelty effect analysis

The choice of Sync Date as the most appropriate anchor for baseline measurement was further supported by a range of user-level case studies (see Box 6 in the Multimedia Appendix for demonstrative examples). The resulting classification of each participant into one of four cohorts based on their syncing behaviour under this baseline definition is shown in Table 3.

**Table 3.** Participant classification based on syncing behaviour.

| Cohort Name | Criterion | Cohort Size (%) |
| --- | --- | --- |
| De Novo Trackers | <10 days of tracking history before Sync Date | 273 (22.1%) |
| Quitters | Last recorded activity <30 days after Sync Date | 84 (6.8%) |
| Fragmented | Any data gap >60 consecutive days post-sync | 79 (6.4%) |
| High-Quality Sync | ≥10 days pre-sync AND consistent post-sync engagement with no gaps >60 days | 797 (64.6%) |
|  |  | Total: 1233 |

Note: n=55 out of the 1,288 participants failed to sync

14-day pre-anchor baseline averages and post-anchor peaks for each cohort × anchor combination are presented in Table 6.1 in the Multimedia Appendix. These results allow a comparison of the three baselines considered in the methodology, and quantification of the novelty effect, indicating the following:

1. Among High-Quality users, the increase from baseline to peak is more pronounced when the baseline is anchored at the Sync Date rather than the Split Date, suggesting a potential novelty effect following app exposure.
2. For De Novo Trackers, using the Sync Date produces an artificially inflated novelty effect. Their observed increase (+4,202 steps) is substantially larger than that of High-Quality users, not because of a stronger behavioural response, but because their baseline is derived from sparse, unreliable pre-sync data.
3. Fragmented users show a similar pattern to High-Quality users across anchors, albeit with a different magnitude.

#### Re-analysis of Mean Daily Step Counts Using Pre-sync Baseline Definition

Under the revised baseline definition, more participants were retained than the restricted per-protocol analysis, with 839 of 1,288 participants included (Figure 6). Baseline characteristics, retention statistics, average daily step patterns, and analyses of seasonal variation are presented in the Multimedia Appendix (Box 8).

**Figure 6.**
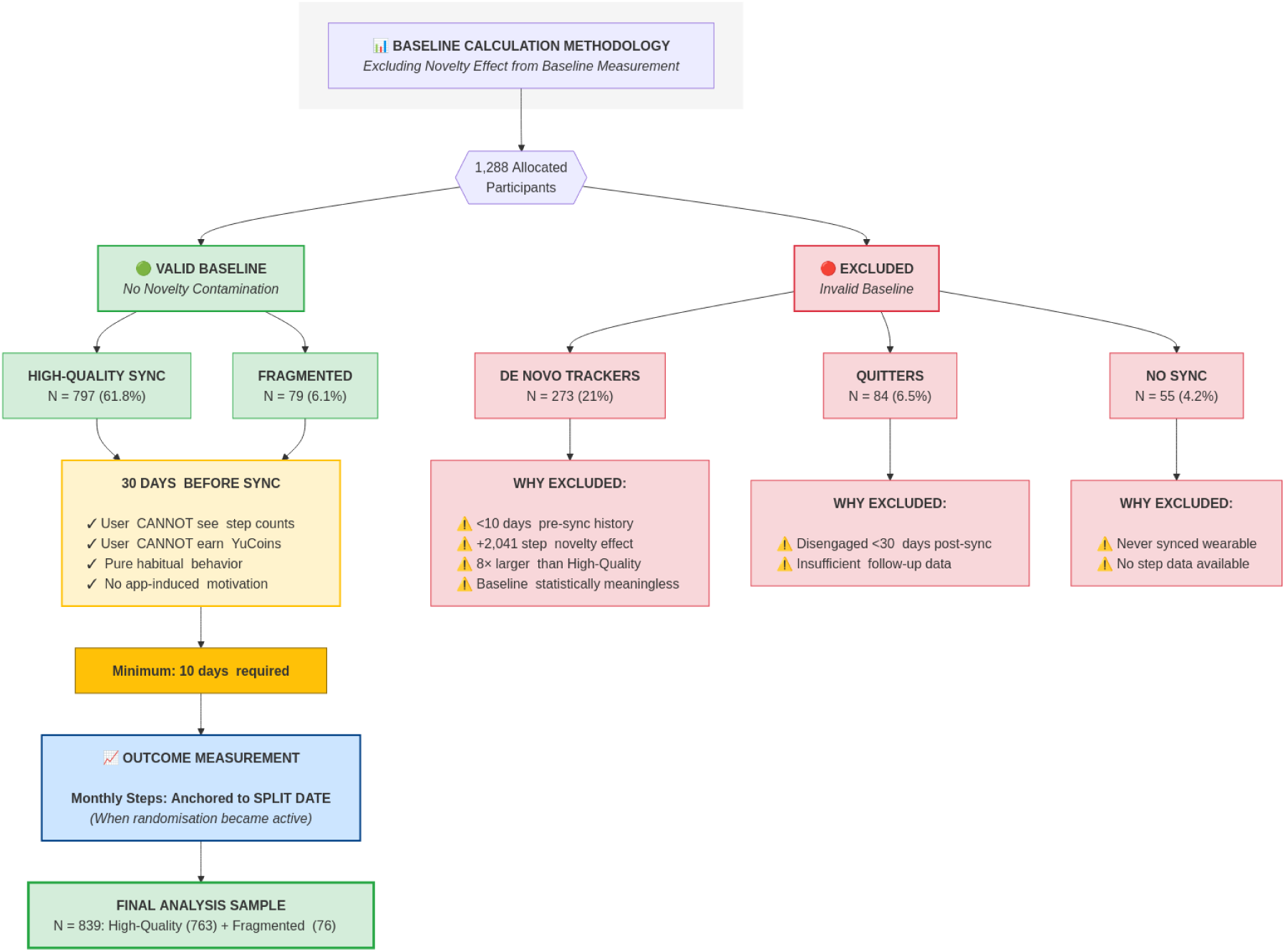
Participant flow for baseline definition, showing included (N=839) and excluded cohorts.

The results showed that the two treatment groups did not differ meaningfully at baseline after adjustment (Intervention group β = −13.22, p = 0.936), indicating balance between groups. In the Control group, mean daily steps did not differ substantively from baseline at any milestone (month 1: β = 44.14, p = 0.749; month 3: β = −6.32, p = 0.966; month 5: β = 40.69, p = 0.794; month 7: β = −81.49, p = 0.605; month 9: β = 220.97, p = 0.144).

Conversely, the time × Intervention interaction terms were positive at every follow-up, indicating higher step counts in the Intervention group relative to Control (month 1: β = 366.18, one-sided p = 0.027; month 3: β = 514.96, one-sided p = 0.005; month 5: β = 533.73, one-sided p = 0.004; month 7: β = 505.42, one-sided p = 0.007; month 9: β = 459.09, one-sided p = 0.014), with all milestones reaching one-sided statistical significance at the 5% level.

As before, baseline-centred steps was a strong predictor of subsequent steps (β = 0.8183, p < 0.001), indicating that participants with higher (alternative) baseline activity tended to record more steps at follow-up. Seasonal effects were again also evident: Autumn (β = −616.12, p < 0.001) and Winter (β = −939.44, p < 0.001), but also Spring (β = −373.32, p < 0.001), were all associated with substantially fewer mean daily steps compared with Summer, all reaching statistical significance. BMI was associated with lower step counts (β = −28.73 per unit increase in BMI, p = 0.018).

In summary, under the alternative baseline definition, the gamified intervention was associated with higher mean daily steps relative to control at all follow-up milestones, with strong evidence of effects throughout the study. Baseline activity, season, and BMI remained important predictors of subsequent step counts. The full model results are provided in the Multimedia Appendix (see Table 8.2).

### Estimating Health Risk Reduction with UK Biobank

The corresponding group-mean trajectory from the predicted partial hazard estimates generated by the LMM model are shown in Figure 7. The fitted model included 3,113 user-timepoint observations contributed by 744 participants (of 839 allocated users with sufficient activity data).

**Figure 7.**
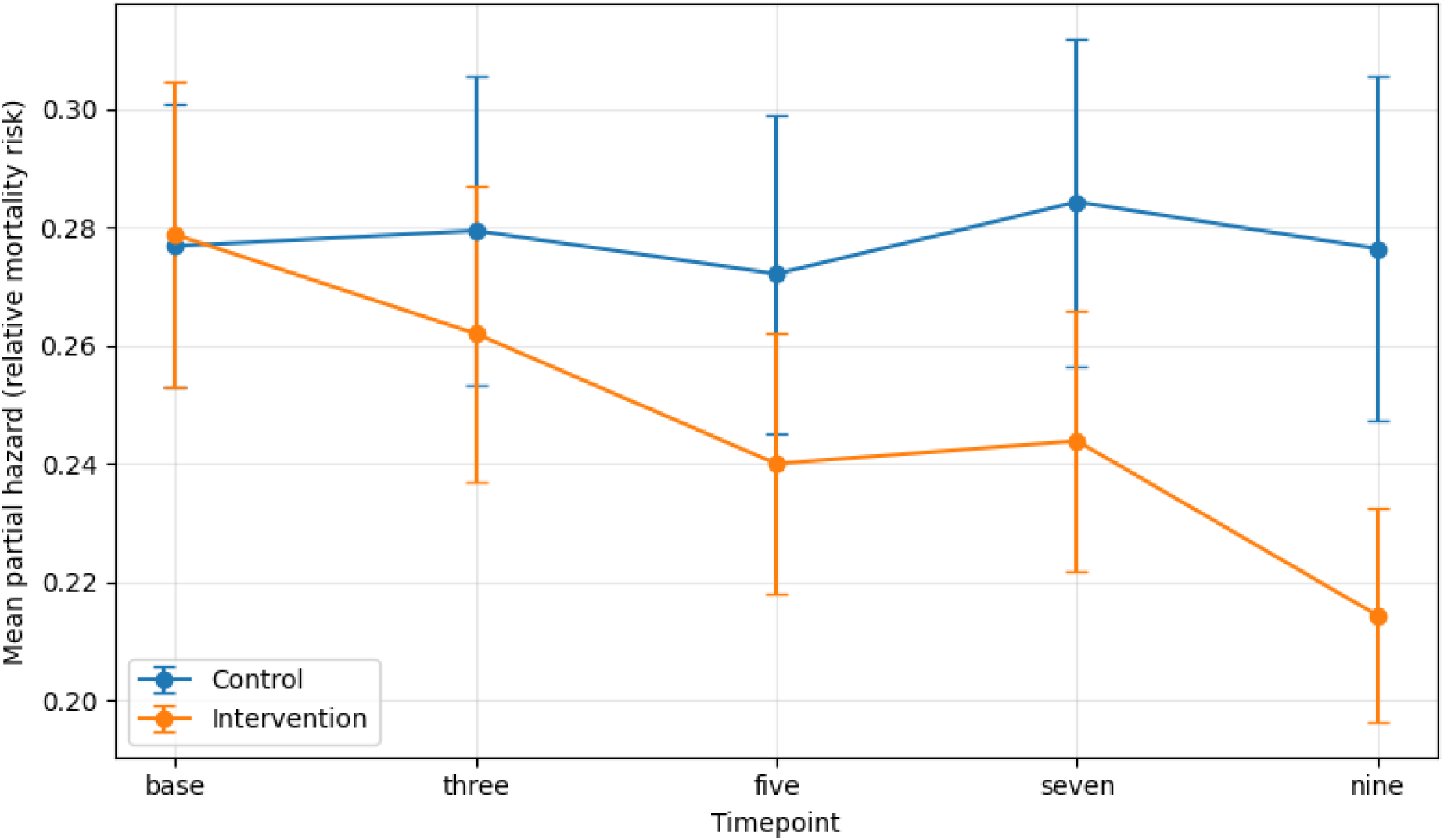
Mean UKB-derived partial hazard by trial timepoint, stratified by treatment arm (n = 839 cohort). Error bars: ±1 SEM.

The results of the LMM were as follows: in the Control group, predicted partial hazard remained essentially flat across follow-up (month 3: β = −0.009, p = 0.121; month 5: β = −0.002, p = 0.687; month 7: β = 0.002, p = 0.718; month 9: β = 0.007, p = 0.261), suggesting that, in the absence of gamification, activity-related risk did not change appreciably over the study period.

The time × Intervention interaction terms were negative at every follow-up point and reached statistical significance at months 5 and 9. The estimated treatment effects were β = −0.007 at month 3 (one-sided p = 0.197), β = −0.018 at month 5 (one-sided p = 0.016), β = −0.009 at month 7 (one-sided p = 0.144), and β = −0.026 at month 9 (one-sided p = 0.002). Translated onto the relative-risk scale, the Intervention arm’s mean partial hazard fell from approximately 6% below Control at month 3 to 12% below at month 5, 14% below at month 7, and 23% below at month 9 (HR ratios 0.94, 0.88, 0.86, and 0.78, respectively). The monotonic widening of this gap, with the largest effect at the latest follow-up, is consistent with a treatment effect that accumulates as users sustain higher activity over the trial, rather than a short-term activation response.

Age and sex were strong predictors of predicted partial hazard, as expected for any UKB-derived mortality model (Age β = 0.029, p < 0.001; Male β = 0.137, p < 0.001), and together with the intercept accounted for the bulk of the explained variance in the LMM model. The residual variation available for treatment-effect detection was therefore modest, which makes the interaction-term significance at months 5 and 9 the more notable. Overall, these results provide evidence that the step-count gains observed in the gamification arm translate into a measurable, progressively widening reduction in predicted all-cause mortality risk on a UKB-validated scale, sustained through the full 9-month study period. The full LMM results as well as the coefficients of the underlying Cox model are provided in Box 7 of the Multimedia Appendix.

## Discussion

### Principal Findings

The primary aim of this study was to evaluate whether the addition of gamification elements to a health and wellbeing app led to positive changes in health behaviour compared with a base version of the app that did not include them. The results demonstrated that, whereas the control group tended to show a small reduction in mean daily steps counts (between ∼200-400 steps on average) across the study’s milestones, the incorporation of gamification elements was instead generally associated with a *positive* increase (from a baseline of roughly 7,800 steps on average), which was generally maintained throughout the duration of the study, peaking at month 5 (with an additional ∼600 steps on average). Similarly, on average, participants who were presented with gamification elements were able to maintain their level of engagement for the duration of the study, compared to the control group which showed a marked decrease of 6-8 days from baseline within the 30 day window over the different milestones. The findings also highlighted a strong seasonal effect for both primary outcomes. In particular, the Summer months were associated with significantly more steps than other months, particularly Autumn and Winter (resulting in differences of 900-1,100 steps) and Spring was associated with increased app engagement (∼2-3 days) compared to Summer. In both outcomes, baseline activity was a strong predictor for subsequent activity. Participants’ BMI was negatively associated with mean daily steps, with each 1-unit increase in BMI associated with a mean step reduction of roughly 60 steps.

Additionally, a sensitivity analysis was conducted to assess the possibility of inflated step counts due to novelty effects occurring during the per-protocol defined baseline period. This uncovered the potential for novelty-effect contamination in the above step analysis, and an alternative baseline was defined post-hoc to investigate its effect. Under this alternative baseline definition, the gamified intervention was again associated with higher mean daily steps relative to control at all follow-up milestones, with strong evidence of effects throughout the study.

Secondary analyses translating step gains to a reduction in mortality risk using outputs obtained via a UKB-derived Cox Proportional Hazards model, showed progressively larger reductions in predicted activity-related hazard in the intervention arm throughout the study (HR ratios ≈0.94→0.78), with significant interaction effects at months 5 and 9. Other secondary outcomes, such as alcohol behaviour, perceived stress, depressive symptoms, generalised anxiety, and work-related outcomes, did not show any statistically significant differences between the two treatment groups over the duration of the trial, except for some short-term reduction in smoking behaviour, but which did not persist in significance following Bonferroni-Holm correction.

### Comparison With Previous Literature

The present findings are broadly consistent with previous evidence suggesting that gamified digital interventions can improve physical activity outcomes, particularly daily step counts. The meta-analysis by Mazeas et al. [18] reported a moderate overall effect of gamification on daily step count, with participants achieving an average increase of approximately 1,610 steps compared with controls (95% CI: 372-2,847). The intervention effects observed in the present study were smaller, with adjusted differences ranging from approximately 330 to 627 additional daily steps across follow-up milestones. However, the larger pooled effect reported in [18] may partly reflect the inclusion of studies that combined gamification with additional behavioural strategies, including financial incentives and other reinforcement mechanisms, rather than isolated gamification alone. In contrast, the present study was specifically designed to evaluate the independent contribution of gamification within a real-world workplace setting while minimising confounding from financial incentives.

The temporal pattern of intervention effects observed in the present study also provides important context regarding the role of novelty effects in digital gamified interventions. Previous literature has frequently reported the largest behavioural improvements immediately after intervention initiation, followed by gradual attenuation over time, often interpreted as a decline in engagement as the novelty of the intervention fades [44]. However, our findings suggest a more complex pattern. Using the original baseline definition, the largest adjusted effect was observed at Month 5 (+627 daily steps), with smaller but still observable differences at Months 7 and 9, indicating that intervention effects were not restricted solely to the earliest stages of engagement. Importantly, sensitivity analyses using a revised baseline definition based on participants’ physical activity prior to first app synchronisation further strengthened this interpretation. By defining the baseline using the 30-day pre-sync period, before participants could view their activity data or interact with app-based rewards, the intervention effects became more stable across follow-up milestones, with adjusted differences ranging from approximately 366 to 534 additional daily steps and statistically significant effects persisting throughout the study. These findings support the presence of an initial behavioural activation or novelty effect occurring immediately after first app exposure in both groups, which may artificially inflate baseline estimates when traditional post-download baseline windows are used. Collectively, these results suggest that gamification-related behavioural changes may be more sustained than previously assumed once early activation effects associated with app onboarding and self-monitoring are appropriately accounted for.

The present study addresses several other limitations repeatedly highlighted in prior literature [12]. Existing evidence has often relied on highly controlled research settings, relatively brief intervention periods, or interventions targeting selected clinical populations [25]. By comparison, this trial evaluated a long-term, gamified intervention embedded within a real-world workplace platform, with everyday user engagement over nine months. In addition, whereas much of the previous literature has focused exclusively on behavioural outcomes such as step count or activity minutes, the present study also explored potential downstream implications for health-risk reduction using UK Biobank-derived modelling approaches, extending the discussion beyond behaviour change alone toward possible long-term health impact.

### Study Limitations

This study has several limitations that could affect interpretation and should be taken into account before generalising to the broader working population. Approximately 67% of the participants invited to the study were excluded from the per-protocol analysis due to non-adherence, protocol deviations, having health limitations, not downloading the app on time, or not faithfully following the study’s instructions and synchronising in a timely fashion. This may impact the generalisability of the results, e.g., by biasing the final analysis towards more engaged users. However, no statistically significant differences were identified in baseline characteristics between the intervention and control groups, reassuring that the exclusion did not introduce substantial bias or imbalance between the two groups. Furthermore, the alternative baseline definition identified through the sensitivity analysis, resulted in a lower number of exclusions (∼35%) and produced similar findings. Nevertheless, it is worth acknowledging that there still exists some possibility that some unmeasured or latent variables exist, which could have led to systematic biases in terms of determining which participants make it into the analysed sample.

Due to practical constraints, it was not possible to do pure block randomisation at a single point in time over all companies, deviating from the originally proposed pre-registration document [30]. However, the two groups were sufficiently close in numbers, and a statistical analysis showed appropriate stratification. Additionally, although models were adjusted for season and company clustering, the staggered enrolment and the open-label, workplace-based nature of the study could have affected the timing and extent of intervention exposure across participants, potentially complicating causal attribution. The open-label design also means participants were aware of their own allocation and thus subject to expectancy or novelty effects that could inflate apparent treatment differences. We accounted for this in the sensitivity analysis, where an alternative baseline period definition was defined on the basis of the exact sync date. This led to an increased sample size of 839 randomised participants, comprising highly engaged users, which confirmed findings similar to the per-protocol analysis.

Questionnaire-based secondary outcomes suffered from substantial attrition, reducing power and increasing uncertainty for mental-health, smoking, alcohol, and other self-reported measures, limiting their usefulness in this context. Similarly, the hazard model estimates risk reductions based on step-count differences at the initial point of risk assessment; however, in reality, it is reasonable to assume that risk reduction would likely depend on sustained increases in step counts rather than on increases at initial conditions alone. And while this study has shown sustained effects at 9 months, this is short relative to the time horizon for observing actual clinical endpoints such as mortality or major morbidity, and thus, actual risk outcomes following long-term sustained increases in physical activity could not be assessed here directly. Despite this, there was some statistical evidence of risk reduction attributable to the gamification treatment within the nine-month window, on the basis of the Cox model associating step increases with a risk reduction, and therefore sustained step increases, even if modest, giving rise to small but progressive risk reduction. It is important to note, however, that while we took care in ensuring that the studied UK Biobank subpopulation used for the creation of the Cox Proportional Hazards model had similar characteristics as our target population (i.e. participants of working age in a company setting), there is still a possibility of systematic differences between the two samples, and therefore absolute mortality reductions should still be interpreted cautiously.

Finally, the gamified app combined many behavioural techniques (levels, leaderboards, streaks, duels, avatars), hence we cannot isolate which specific elements drove the observed effects. We also emphasise that the primary analyses presented are per-protocol; the nature of the study prevented us from meaningfully comparing against intention-to-treat estimates (which would include non-synchronisers and reflect assignment rather than adherence). One might expect that an intention-to-treat analysis would produce smaller effect sizes; however, we point out that the MICE-imputed analysis yielded findings similar to the per-protocol analysis in this instance.

### Implications for Practice

The trial provides evidence that gamification (beyond equivalent monetary rewards) can increase physical activity and sustain higher app-engagement in a real-world employed population, with the largest modelled effect observed around the mid-point of the study (but sustained throughout the study under the alternative baseline considered in the sensitivity analysis). Although the individual-level increases in daily steps are modest, they are meaningful when considered across populations reached via employer programmes; even relatively small, sustained average increases could shift cohort risk profiles and lead to better outcomes for both the work population in question as well as the employer. This finding suggests two practical implications for health-risk managers and employers: first, gamified elements can be an effective, low-friction tool to boost engagement and activity, particularly in the short- to medium-term, supporting workplace prevention and wellbeing objectives.

Second, converting such behaviour change into measurable reductions in clinical risk or mortality likely requires larger or more sustained behavioural shifts, longer follow-up, or integration with additional clinical supports to influence hard outcomes and claims costs. This means that gamification strategies could be an effective component of a larger risk-management framework, especially for providing short- to medium-term incentives for targeted interventions aimed at behavioural change, ensuring long-term engagement as necessary to ensure appropriate health risk reduction.

### Future Research

Given that interpretation and clinical impact insights are limited by the study’s duration and pragmatic design, future research should focus on longer follow-up and intention-to-treat analyses to evaluate the persistence of behaviour change and its relation to clinical endpoints within the same study population. This could be further combined with structured clinical and/or wellbeing support (for example, exercise programmes or coaching) to determine whether larger, clinically meaningful risk reductions can be achieved. If the timelines required to observe meaningful clinical endpoints are infeasible in study contexts, observing near-term biomarkers as proxies for long-term risk may be a more feasible approach.

Additionally, future work could focus on identifying which gamification features (for example, social competition, narrative progression, or streaks) are most effective and for which subgroups, and to what extent there is potential for personalisation.

Finally, it would be useful to examine organisational factors that influence engagement with such programs and potential heterogeneity of benefit across workforce subgroups, as well as any potential unintended consequences, such as feelings of coercion due to perceptions (or actual risks) of negative effects from non-compliance in relation to their employment, or other adverse behaviours such as attempts to ‘game’ the system, or competition-related stress.

## Conclusion

This 9-month RCT involving participants in a workplace setting demonstrated that adding gamification elements to a health and wellbeing app intended to promote positive health behaviours (with a focus on physical activity, measured by mean daily step counts) led to sustained positive change over the study period. By mapping onto a mortality risk prediction model trained on UK-biobank data, the study also demonstrated progressive risk reduction over the study period, indicating that, if maintained, the observed behaviour change could meaningfully improve long-term health outcomes. Additionally, higher engagement with the app was also observed, which could provide opportunities for greater exposure to, and interaction with a wider range of health behaviour-promoting interventions. We conclude that gamification appears valuable as a scalable engagement strategy for workplace health promotion, particularly for targeted short- to medium-term behavioral change interventions operating within a larger risk-management framework.

## Data Availability

All data produced in the present study are available upon reasonable request to the authors

## Funding

This work was supported by a Knowledge Transfer Partnership (KTP) grant from Innovate UK awarded to the University of Essex and YuLife Ltd (grant 10059363; KTP reference 13536), which facilitates collaboration between industry and academic institutions. Four authors (JR, MR, BF, and MD) are employed by the industry partner, YuLife Ltd. The manuscript was drafted entirely by the academic team; JR, MR, BF, and MD contributed to content review, independent verification of results, and provided supervisory and administrative support within the broader project. The academic authors declare no financial interests related to YuLife Ltd beyond the scope of the KTP grant.

## Ethical Considerations

The study was approved by the Institutional Review Board/Ethics Committee of the University of Essex (Application number: ETH2324-0764). All participants provided informed consent in accordance with institutional guidelines and the Declaration of Helsinki.

## Notes

### Clinical Trial

https://osf.io/926pd

### Clinical Protocols

https://osf.io/926pd

