## Appendix for "Impact of Gamification in Behaviour Change Intervention: A Randomised Controlled Trial with YuLife’s Health and Wellbeing App"

**This is an Appendix to a full manuscript published in Medarxiv, entitled “Impact of Gamification in Behaviour Change Intervention: A Randomised Controlled Trial with YuLife's Health and Wellbeing App” by A. Salami et al.**

Table of contents

**Box 1. Features of the YuLife app**

**Box 2. LMM results – Per-protocol analysis**

Mean daily step activity

App engagement

**Box 3: Effect of seasonality**

**Box 4: Step count analysis using multiple imputation**

**Box 5: Secondary outcomes:**

Effects on sleep duration

Effects on smoking

Effects on the frequency of alcohol consumption

Effects on perceived stress (as measured via the PSS4 scale)

Effects on generalised anxiety (as measured via the GAD2 scale)

Effects on depression (as measured via the PHQ2 scale)

Effects on work-related outcomes

Collective results following Bonferroni-Holm correction

**Box 6: Why Milestone Choice Matters**

User-Level Case Studies

Novelty effect spike analysis

**Box 7: Estimating Health Risk Reduction from steps via UK Biobank**

Characteristics of the trained Cox Proportional Hazards model

Mapping participant steps to partial hazard using the trained Cox model

**Box 8: LMM results – Alternative baseline definition analyses.**

**Box 9 - Questionnaires used in the study**

Baseline Health Questionnaire

Follow-up questionnaires

Dynamic Health Questionnaire

**References**

### Box 1. Features of the YuLife app

The YuLife app features a variety of components which incorporate gamification techniques. Gamification techniques may relate to one or more Behavioural Change Techniques (BCT) – see [[1]](https://paperpile.com/c/PiWPX3/RrE3). Table 1.1 lists some of those more prominent app components below, with a descriptive summary and BCT categories for each one

**Table 1.1:** YuLife app gamification components

| 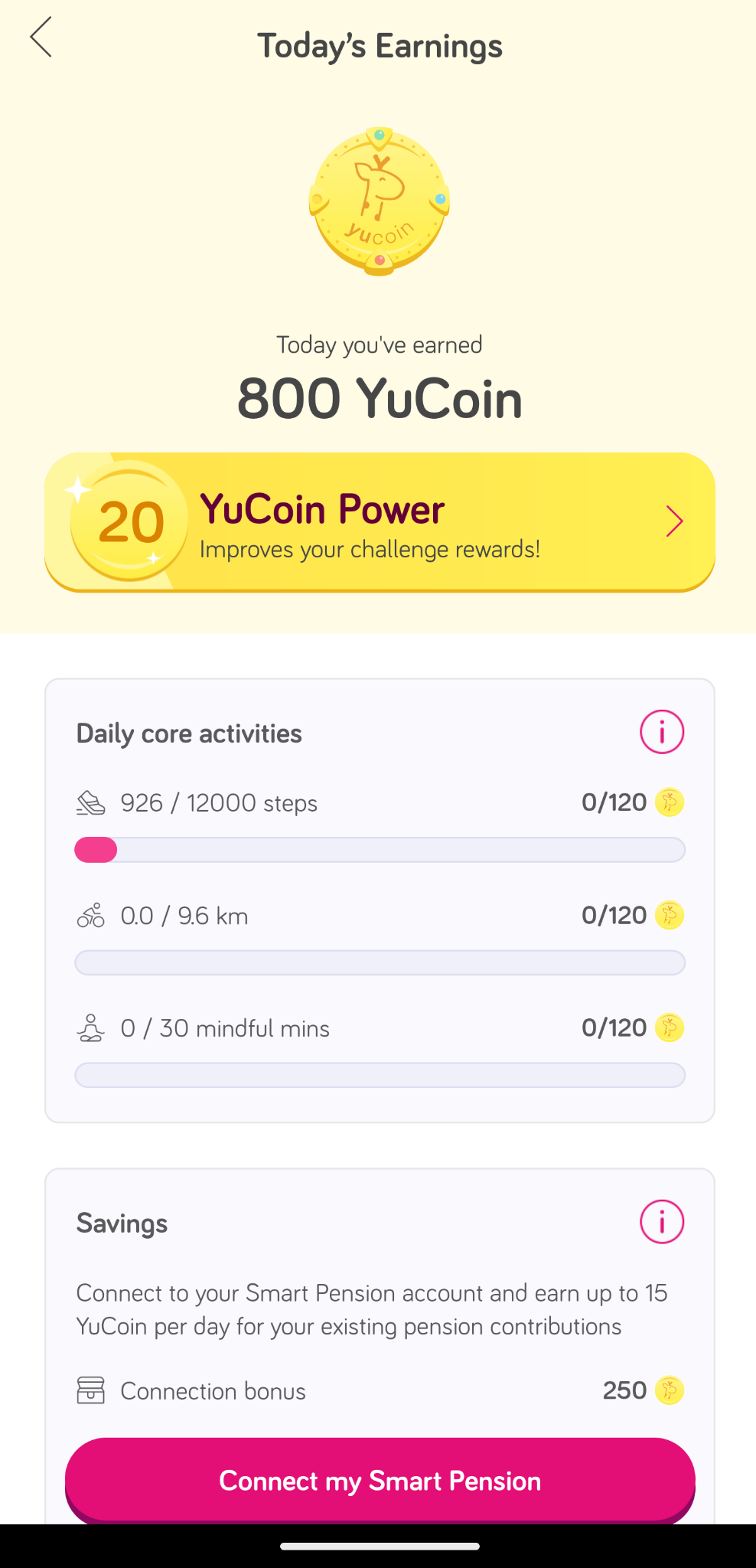 | **YuCoin**: a virtual currency earned through activities such as walking, meditation, and completing challenges, redeemable for vouchers, wellbeing products, or charitable donations.  Relevant BCTs:   - Reward and threat   - Material incentive   - Material reward   - Non-specific reward |
| --- | --- |
| 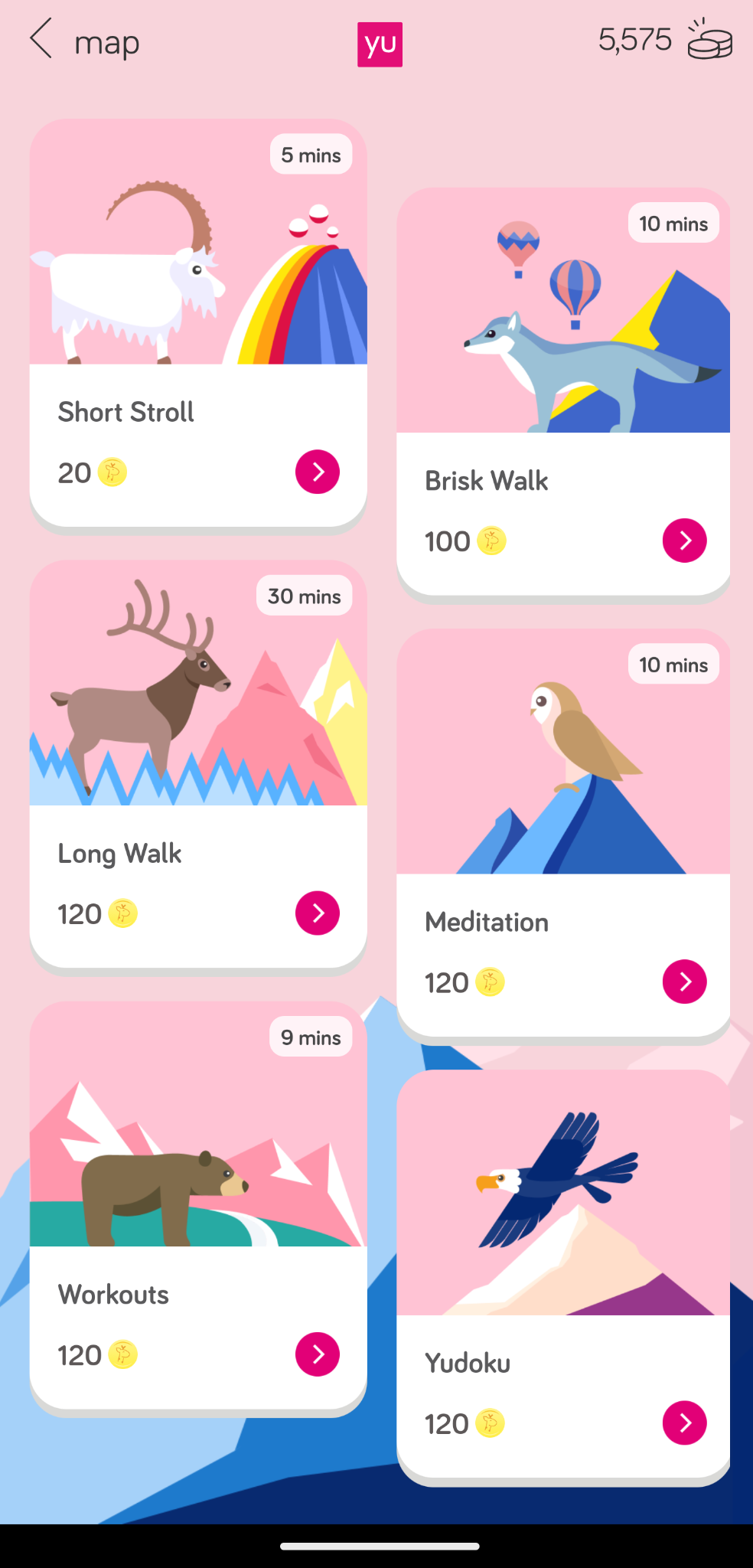 | **Challenges**: task-based activities like walking 1,000 steps in 10 minutes or completing a 10-minute meditation.  Relevant BCTs:   - Goals and planning   - Goal setting (behaviour)   - Problem solving   - Goal setting (outcome)   - Action planning   - Behavioral contract   - Commitment - Feedback and monitoring   - Feedback on behaviour   - Self-monitoring of behaviour   - Biofeedback - Shaping knowledge   - Instruction on how to perform the behaviour - Repetition and substitution   - Habit formation - Reward and threat   - Material incentive (behaviour)   - Self-incentive - Self-belief   - Focus on past success |
| 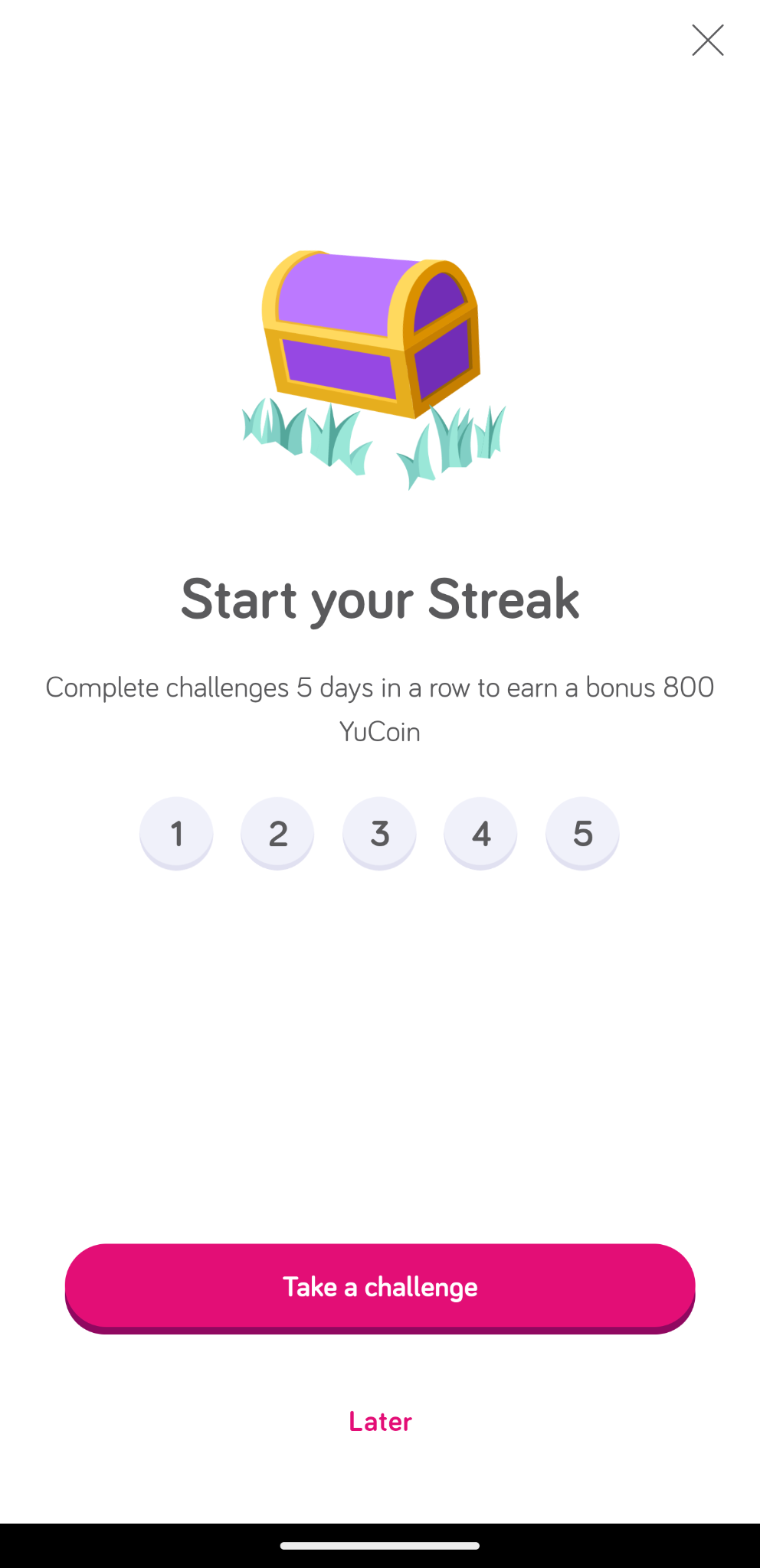 | **Streaks**: rewarded users for consecutive days of activity or challenge participation.  Relevant BCTs:   - Goals and planning   - Goal setting (behaviour)   - Goal setting (outcome)   - Behavioral contract   - Commitment - Feedback and monitoring   - Self-monitoring of behaviour - Natural consequences   - Anticipated regret - Repetition and substitution   - Behavioural practice   - Habit formation   - Generalisation of target behaviour - Reward and threat   - Material incentive (behaviour)   - Self-incentive |
| 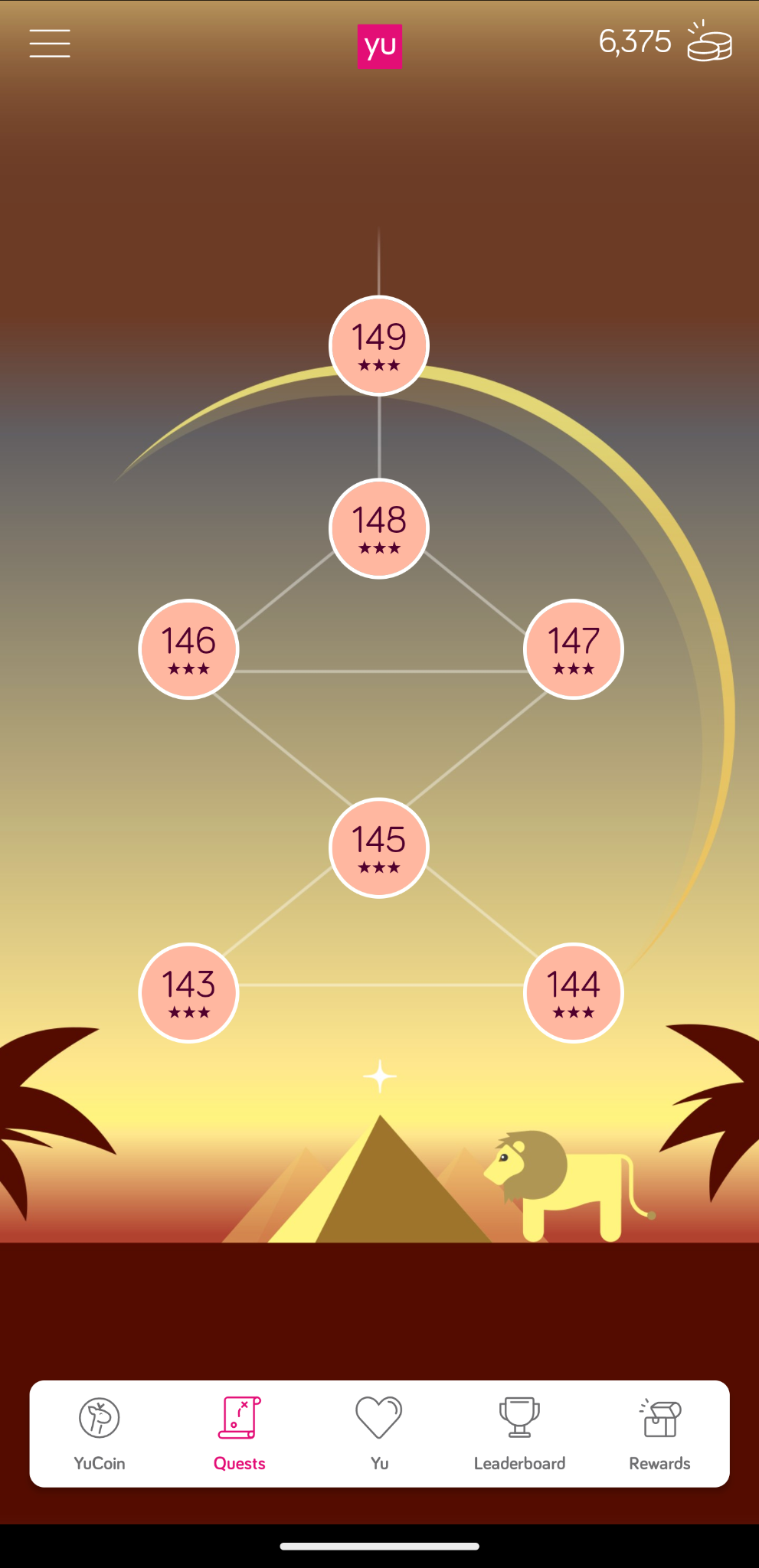 | **Levels**: users unlocked additional challenges as they progressed.  Relevant BCTs:   - Comparison of behaviour   - Social comparison - Reward and threat   - Non-specific reward - Identity   - Framing / reframing   - Identity associated with changed behaviour - Scheduled consequences   - Situation-specific reward - Covert learning   - Vicarious consequences |
| 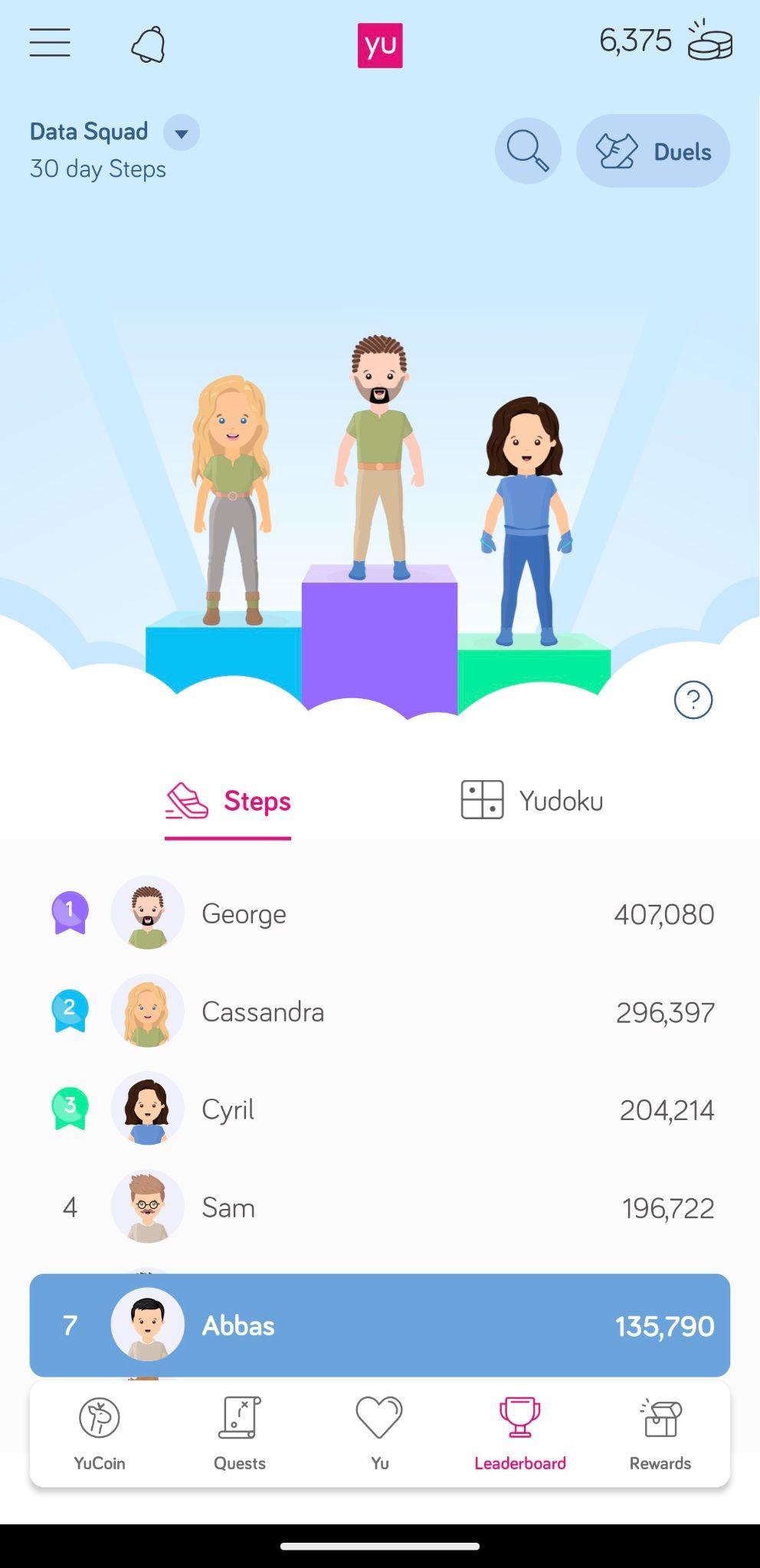 | **Leaderboards**: displayed users' 30-day average steps in comparison to peers.  Relevant BCTs:   - Comparison of behaviour   - Social comparison - Reward and threat   - Social incentive |
| 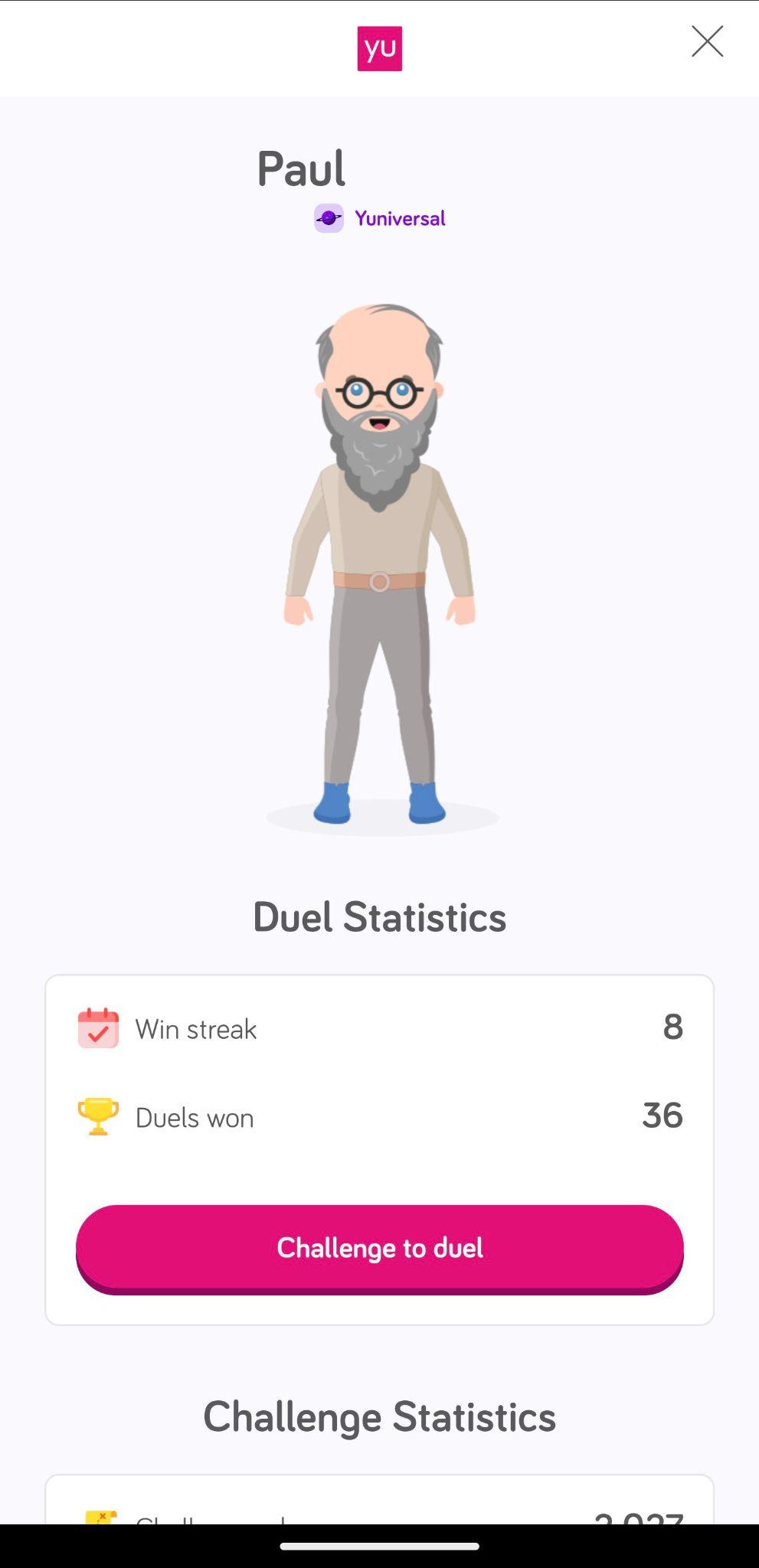 | **Duels**: allowed users to compete directly with colleagues on step counts over 24-hour windows, with the option to wager YuCoin on the outcome, adding a competitive and incentive-driven element to physical activity.  Relevant BCTs:   - Social support   - Social support (unspecified) - Comparison of behaviour   - Social comparison   - Demonstration of the behaviour |
| 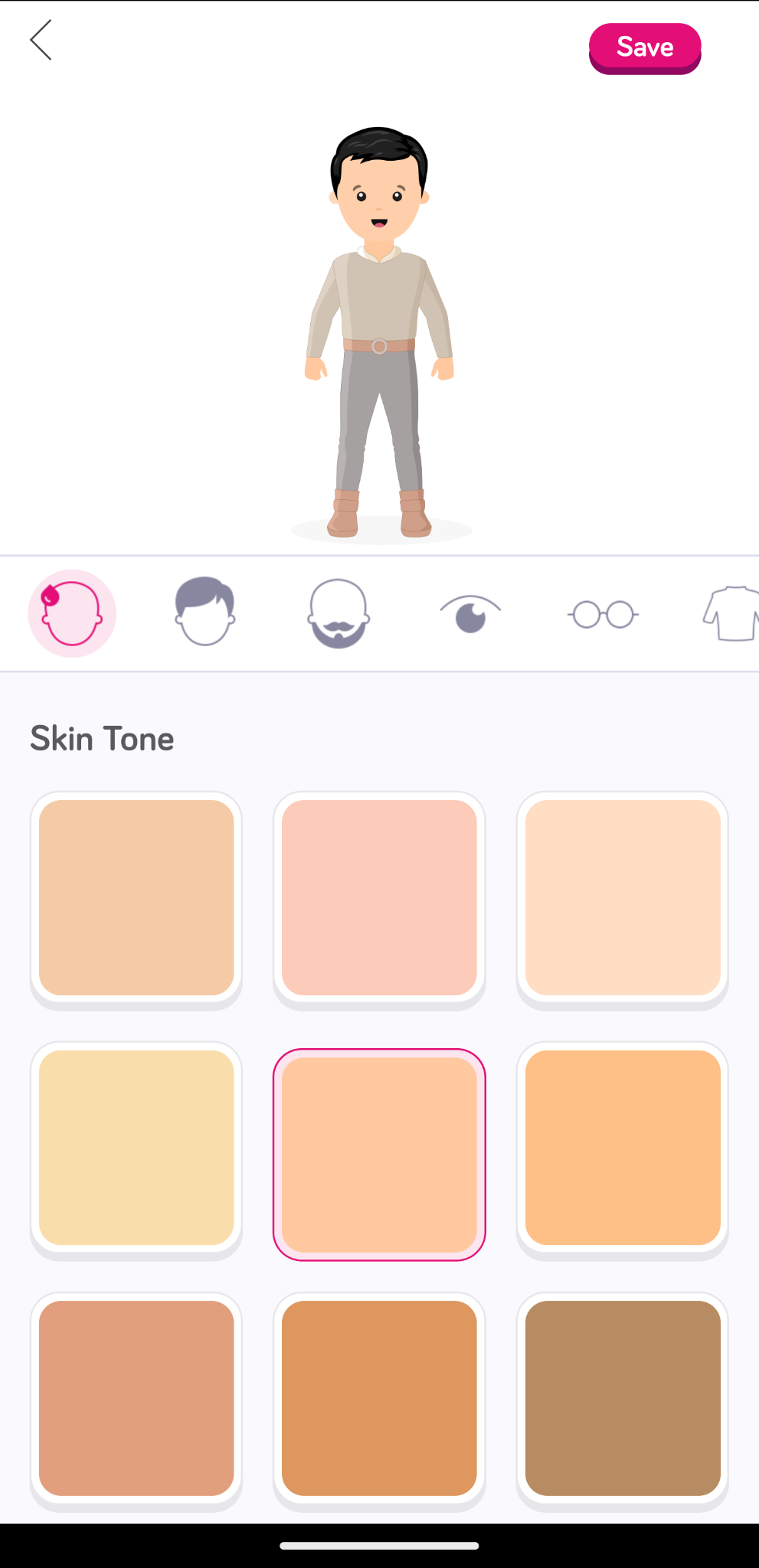 | **Avatars and Virtual Worlds**: users could customise avatars and earn visual achievements (e.g., clothing or temporary boosts such as double YuCoin rewards for completing challenges) that unlocked different "universes" within the app as a narrative of progress.  Relevant BCTs:   - Identity   - Valued self-identity   - Identity associated with changed behaviour |
| 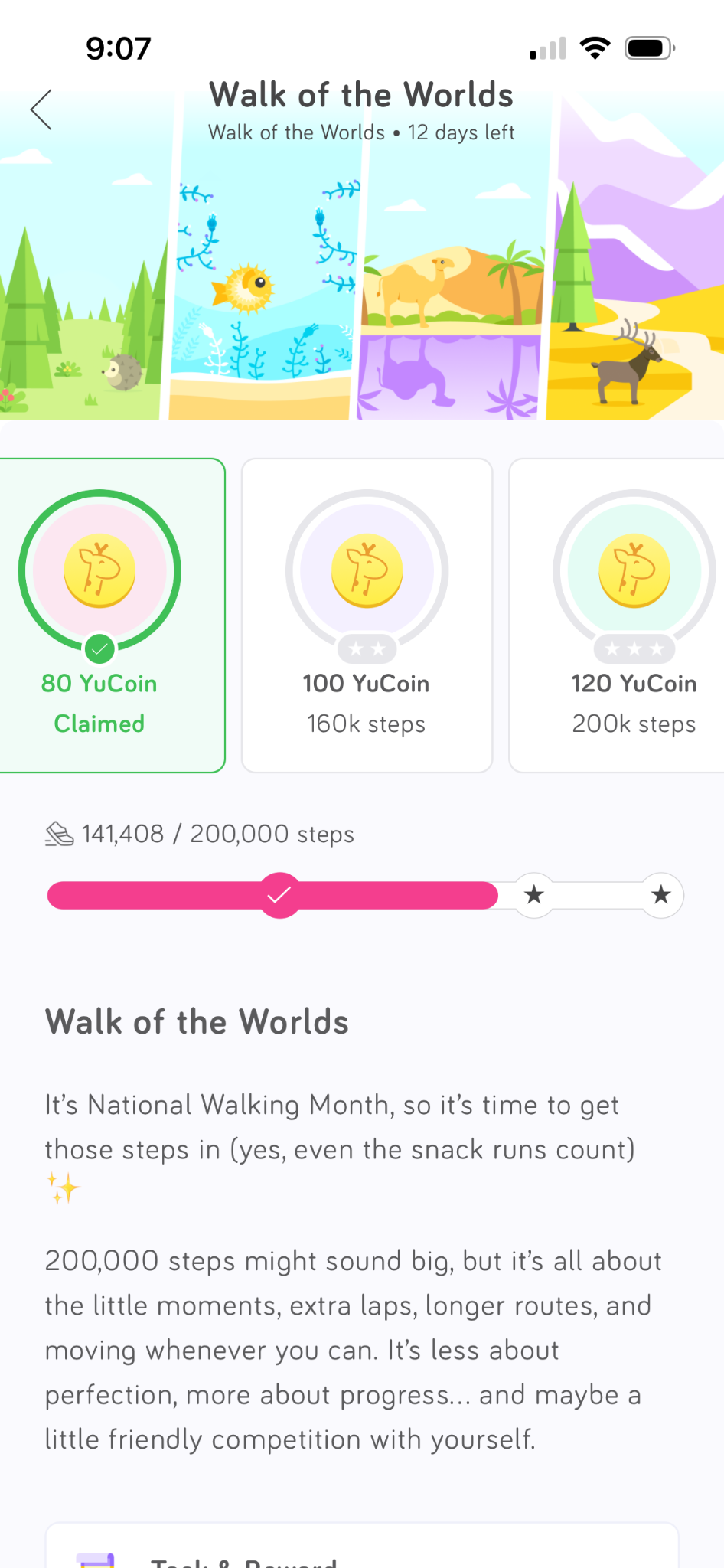 | **Goals and Events**: broader time-limited objectives (e.g., 180,000 steps in a month) provided additional incentives.  Relevant BCTs:   - Goals and planning   - Goal setting (behaviour) - Associations   - Prompts/cues   - Exposure - Reward and threat   - Material incentive (behaviour) |
| 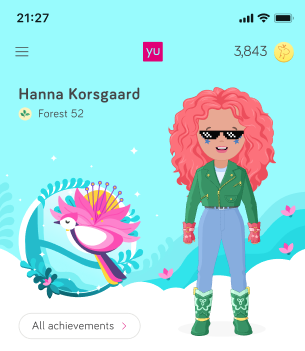 | **Badges**: in-app symbols of achievement and progression (such as diamonds on the app logo).  Relevant BCTs:   - Reward and threat   - Non-specific reward   - Social incentive |

### Box 2. LMM results – Per-protocol analysis

Tables 2.1 and 2.2 below present the results of the Linear Mixed Model (LMM) analysis of mean daily step counts and app engagement, respectively, for the per-protocol analysis.

#### Mean daily step activity

Table 2.1 shows the mean daily step count results. This was produced by fitting an LMM model on the strict per-protocol cohort after listwise drop on Age, Gender, BMI, and Steps, using the following R formula:

STEPS ~ Time * Group
+ Baseline_Centered
+ Season
+ Gender
+ Age_Group
+ BMI
+ (1 | Business_Name / YuLife_User_ID)

**Table 2.1:** Linear mixed-effects model: Mean daily step activity

**Fixed effects:**

| **Effect** | **Estimate** | **Std.Err.** | **P>\|z\|** | **One-sided p-value** |
| --- | --- | --- | --- | --- |
| (Intercept) | 8852.75 | 1,019.88 | <0.001 |  |
| Milestone: Month 1 | -280.878 | 187.864 | 0.135 |  |
| Milestone: Month 3 | -368.658 | 196.111 | 0.06 |  |
| Milestone: Month 5 | -194.997 | 225.735 | 0.388 |  |
| Milestone: Month 7 | -237.366 | 222.062 | 0.285 |  |
| Milestone: Month 9 | -197.569 | 204.819 | 0.335 |  |
| Group: Intervention | -31.784 | 197.989 | 0.872 |  |
| Baseline (centred) | 0.8531 | 0.0193 | <0.001 |  |
| Season: Spring | -194.758 | 141.363 | 0.168 |  |
| Season: Autumn | -943.503 | 166.368 | <0.001 |  |
| Season: Winter | -1,145.45 | 165.09 | <0.001 |  |
| BMI | -60.836 | 13.301 | <0.001 |  |
| Month 1 × Intervention | 341.979 | 232.765 | 0.142 | 0.071 |
| Month 3 × Intervention | 473.844 | 244.362 | 0.053 | 0.027 |
| Month 5 × Intervention | 626.542 | 246.466 | 0.011 | 0.006 |
| Month 7 × Intervention | 330.641 | 251.786 | 0.189 | 0.095 |
| Month 9 × Intervention | 480.914 | 260.805 | 0.065 | 0.033 |

Note: Reference categories are: Milestone = Baseline (Month 0), Group = Control, Season = Summer. Gender and age group were included as covariates but not shown here for brevity.

#### App engagement

Table 2.2 shows app engagement results. This was produced using the following R formula:

ENGAGEMENT_PCT ~ Time * Group

+ Baseline_Centered

+ Season

+ (1 | Business_Name / YuLife_User_ID)

**Table 2.2:** Linear mixed-effects model: App engagement

| **Effect** | **Estimate** | **SE** | **P>\|z\|** | **One-sided p-value** |
| --- | --- | --- | --- | --- |
| (Intercept) | 0.491 | 0.032 | <0.001 |  |
| Milestone: Month 3 | −0.218 | 0.021 | <0.001 |  |
| Milestone: Month 5 | −0.212 | 0.024 | <0.001 |  |
| Milestone: Month 7 | −0.201 | 0.024 | <0.001 |  |
| Milestone: Month 9 | −0.255 | 0.021 | <0.001 |  |
| Group: Intervention | 0.018 | 0.029 | 0.531 |  |
| Baseline (Centred) | 0.732 | 0.043 | <0.001 |  |
| Season: Spring | 0.075 | 0.016 | <0.001 |  |
| Season: Autumn | −0.019 | 0.017 | 0.268 |  |
| Season: Winter | −0.016 | 0.019 | 0.378 |  |
| Month 3 × Intervention | 0.205 | 0.027 | <0.001 | <0.001 |
| Month 5 × Intervention | 0.182 | 0.027 | <0.001 | <0.001 |
| Month 7 × Intervention | 0.170 | 0.027 | <0.001 | <0.001 |
| Month 9 × Intervention | 0.175 | 0.027 | <0.001 | <0.001 |

Note: Reference categories are baseline engagement, control group, and Summer season.

### Box 3: Effect of seasonality

The effect of season is also notable in the model results. With Summer serving as the reference season, all other seasons demonstrate fewer average steps, with Autumn (−944 steps, p<0.001) and Winter (−1145 steps, p<0.001) reaching statistical significance. Figure 3.1 shows the distribution of steps by season, confirming the model findings that steps increase in Summer relative to other seasons.

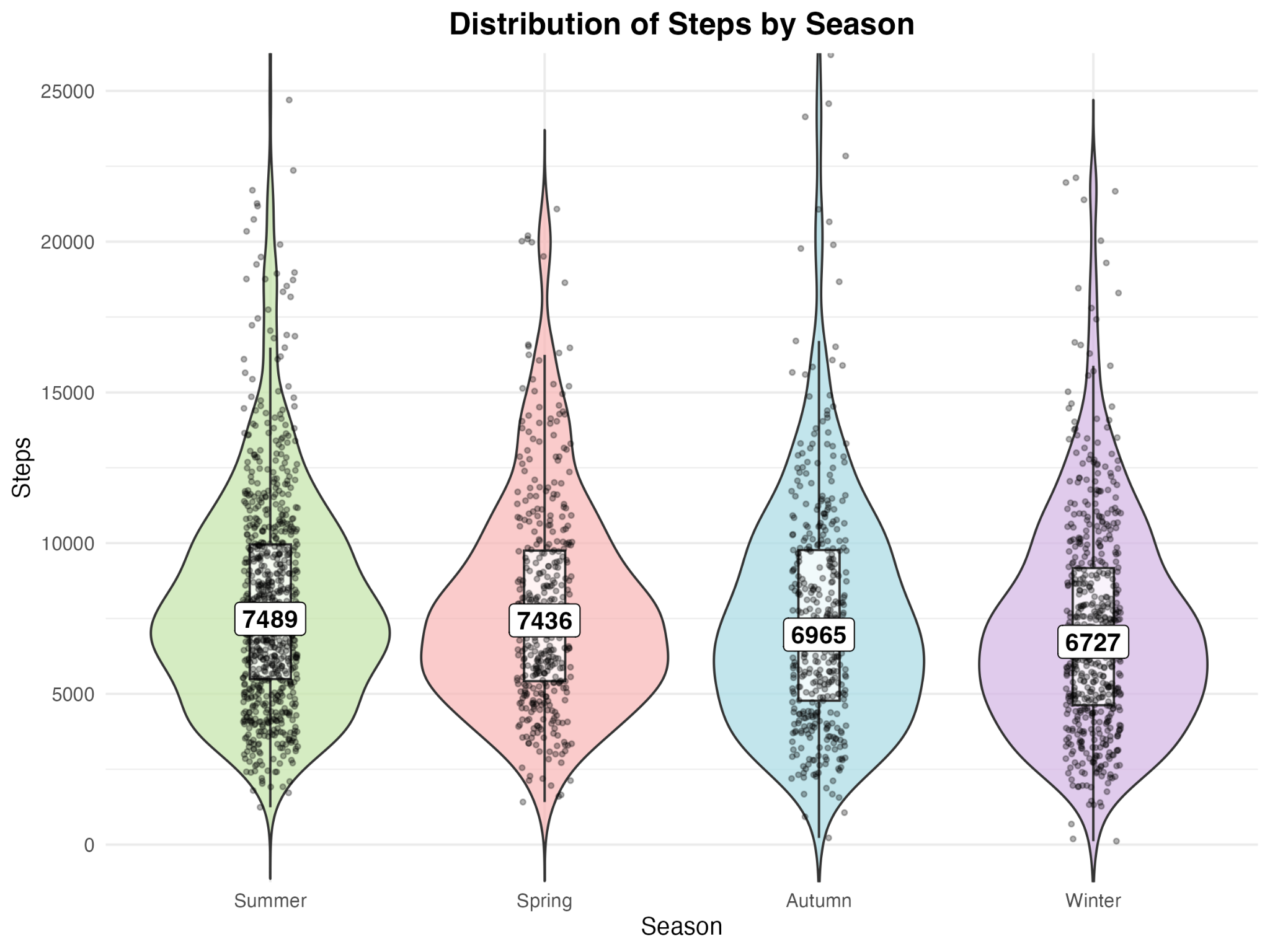

**Figure 3.1:** Distribution of steps by season: While the distributions are similar in shape, Summer tends to have more steps (median 7489), followed by Spring (7436), Autumn (6965), and Winter (6727).

### Box 4: Step count analysis using multiple imputation

The strict per-protocol analysis in the main paper excluded participants with insufficient baseline or follow-up steps. Separate from the above analysis, we conducted a modified intention-to-treat analysis by performing multiple imputation to account for the missing steps, as discussed in the “Data Handling” section of the main manuscript, and re-running the analysis as per the per-protocol criteria. Table 4.1 below shows the result of this analysis.

**Table 4.1:** Multiple imputation results (over 50 iterations of the MICE algorithm)

| **Effect** | **Estimate** | **Std.Err.** | **P>\|z\|** | **One-sided p-value** |
| --- | --- | --- | --- | --- |
| (Intercept) | 7195.52 | 177.955 | < 0.001 |  |
| Month 3 | 137.011 | 128.731 | 0.287 |  |
| Month 6 | -56.158 | 255.298 | 0.826 |  |
| Month 9 | 281.852 | 422.377 | 0.508 |  |
| Intervention group | -0.1212 | 151.565 | 0.999 |  |
| Baseline steps (centred) | 0.8539 | 0.0161 | < 0.001 |  |
| Spring | 169.483 | 185.245 | 0.36 |  |
| Winter | 595.217 | 486.082 | 0.226 |  |
| Month 3 × Intervention group | 371.851 | 183.552 | 0.043 | 0.022 |
| Month 6 × Intervention group | 341.635 | 190.512 | 0.073 | 0.037 |
| Month 9 × Intervention group | 268.369 | 201.908 | 0.184 | 0.092 |

The results above largely agree with the original analysis; the intervention is associated with a positive effect in step counts, of the order of ~300-400 steps, achieving statistical significance in months 3 and 6.

### Box 5: Secondary outcomes:

This section reports the secondary outcomes considered in this study, namely:

- Effects on sleep
- Effects on smoking
- Effects on the frequency of alcohol consumption
- Effects on perceived stress (as measured via the PSS4 scale)
- Effects on generalised anxiety (as measured via the GAD2 scale)
- Effects on depression (as measured via the PHQ2 scale)
- Effects on work-related outcomes

All analyses are performed on the same sample of participants (n=427) as in the main study, using Wilcoxon and Mann-Whitney tests for within- and between-group analyses, respectively, as described below.

In summary, while the occasional isolated significant effect was observed, when considered as a family of tests with a Bonferroni-Holm correction applied, no significant effects were observed in either the Wilcoxon or Mann-Whitney test for any outcome.

#### Effects on sleep duration

Effects on sleep duration were assessed by categorising numerical values provided in the baseline and milestone health questionnaires (HQ) into three bins: "short_sleep (≤6h)", "recommended_sleep (6-9h)", and "long_sleep (>9h)". For users without an HQ response, equivalent responses from the Dynamic Health Questionnaire (DHQ) were used instead (see Box 8 for details on the questionnaires). ‘Recommended sleep’ was then mapped to a score of 1, with the other two categories mapped to 0.

A Wilcoxon Signed Ranks test was performed on the per-participant score differences (i.e. score at follow-up – score at baseline) on both treatment groups. Additionally, a Mann-Whitney U test was performed to compare the two score-difference distributions between the groups. No significant improvement in ideal sleep was observed at any milestone in either group, and no improvement was observed in the intervention group relative to the control group. Table 5.1 summarises the results of this analysis.

**Table 5.1:** Sleep duration analysis. Reported p-values are for one-sided hypotheses (corresponding to an improvement, i.e. an increase in the binary sleep score).

| **Milestone** | **No. Observations (Control)** | **No. Observations (Intervention)** | **Median score difference (Control)** | **Median score difference (Intervention)** | **Wilcoxon p-value (Control)** | **Wilcoxon p-value (Intervention)** | **Mann-Whitney p-value** |
| --- | --- | --- | --- | --- | --- | --- | --- |
| Month 3 | 98 | 101 | 0 | 0 | 0.1526 | 0.3089 | 0.706 |
| Month 6 | 78 | 88 | 0 | 0 | 0.0934 | 0.4182 | 0.8112 |
| Month 9 | 93 | 92 | 0 | 0 | 0.1526 | 0.406 | 0.7552 |

#### Effects on smoking

Smoking was assessed via a 5-point ordinal scale (see item 6 in the baseline HQ in Box 9), binarised such that ‘No’ was given a score of 0, and all other categories were given a score of 1. Wilcoxon Signed-Rank tests were performed on per-participant score differences for both treatment groups, and a Mann-Whitney U test was performed to compare the two score-difference distributions between the groups, as above. While no significant decrease in smoking behaviour was observed *within* groups at any milestone, the Mann-Whitney U test was significant at milestones 3 and 6, indicating a potential difference in smoking behaviour between the two groups. Table 5.2 summarises the results of this analysis.

**Table 5.2:** Analysis of change in smoking habit. Reported p-values are for one-sided hypotheses (corresponding to a reduction in the severity of smoking habits). ‘NaN’ values indicated that there was no change in score between baseline and follow-up for any of the participants.

| **Milestone** | **No. Observations (Control)** | **No. Observations (Intervention)** | **Median score difference (Control)** | **Median score difference (Intervention)** | **Wilcoxon p-value (Control)** | **Wilcoxon p-value (Intervention)** | **Mann-Whitney p-value** |
| --- | --- | --- | --- | --- | --- | --- | --- |
| Month 3 | 96 | 99 | 0 | 0 | 0.9861 | nan | 0.0206 |
| Month 6 | 77 | 85 | 0 | 0 | 0.9861 | 0.5 | 0.0106 |
| Month 9 | 90 | 90 | 0 | 0 | nan | 0.9772 | 0.844 |

#### Effects on the frequency of alcohol consumption

Frequency of alcohol consumption was assessed via a 6-point ordinal scale (see item 7 in the baseline HQ in Box 9). Wilcoxon and Mann-Whitney one-sided tests were conducted as above. Significant effects were observed within the control group at all milestones, and in the intervention group at milestone 9. However, no significant difference in the reduction in alcohol frequency was detected between groups. Table 5.3 summarises the results of this analysis.

**Table 5.3:** Analysis of change in frequency of alcohol consumption. Reported p-values are for one-sided hypotheses (corresponding to a reduction in alcohol consumption frequency)

| **Milestone** | **No. Observations (Control)** | **No. Observations (Intervention)** | **Median score difference (Control)** | **Median score difference (Intervention)** | **Wilcoxon p-value (Control)** | **Wilcoxon p-value (Intervention)** | **Mann-Whitney p-value** |
| --- | --- | --- | --- | --- | --- | --- | --- |
| Month 3 | 96 | 99 | 0 | 0 | 0.009 | 0.2499 | 0.8731 |
| Month 6 | 77 | 85 | 0 | 0 | 0.0124 | 0.1527 | 0.9242 |
| Month 9 | 90 | 90 | 0 | 0 | 0.0107 | 0.0329 | 0.7407 |

#### Effects on perceived stress (as measured via the PSS4 scale)

Perceived stress was assessed via the 4-point Perceived Stress Scale (PSS4) [[2]](https://paperpile.com/c/PiWPX3/2o90) (see items 13-16 in the baseline HQ in Box 9). Wilcoxon and Mann-Whitney one-sided tests were conducted as above. Significant effects were observed within the control group at milestone 9, and in the intervention group at milestones 3 and 9. Milestone 3 also showed a significant difference in scores between groups, but this was not sustained through milestones 6 and 9. Table 5.4 summarises the results of this analysis.

**Table 5.4:** Analysis of change in perceived stress. Reported p-values are for one-sided hypotheses (corresponding to a reduction in perceived stress).

| **Milestone** | **No. Observations (Control)** | **No. Observations (Intervention)** | **Median score difference (Control)** | **Median score difference (Intervention)** | **Wilcoxon p-value (Control)** | **Wilcoxon p-value (Intervention)** | **Mann-Whitney p-value** |
| --- | --- | --- | --- | --- | --- | --- | --- |
| Month 3 | 96 | 99 | 0 | -1 | 0.4167 | 0.0136 | 0.06 |
| Month 6 | 77 | 85 | 0 | 0 | 0.2095 | 0.1522 | 0.5047 |
| Month 9 | 90 | 90 | -1 | 0 | 0.0112 | 0.0469 | 0.7562 |

#### Effects on generalised anxiety (as measured via the GAD2 scale)

Generalised anxiety was assessed via the 2-point Generalised Anxiety Disorder scale (GAD-2) [[3]](https://paperpile.com/c/PiWPX3/fRxf) (see items 19-20 in the baseline HQ in Box 9). Wilcoxon and Mann-Whitney one-sided tests were conducted as above. No statistically significant effects were observed in the control group; a statistically significant reduction was observed in the intervention group at milestone 3, but it was not sustained across the remaining milestones. No significant difference indicating a reduction in anxiety was observed between the two groups. Table 5.5 summarises the results of this analysis.

**Table 5.5:** Analysis of generalised anxiety. Reported p-values are for one-sided hypotheses (corresponding to a decrease in generalised anxiety).

| **Milestone** | **No. Observations (Control)** | **No. Observations (Intervention)** | **Median score difference (Control)** | **Median score difference (Intervention)** | **Wilcoxon p-value (Control)** | **Wilcoxon p-value (Intervention)** | **Mann-Whitney p-value** |
| --- | --- | --- | --- | --- | --- | --- | --- |
| Month 3 | 96 | 99 | 0 | 0 | 0.7409 | 0.0475 | 0.075 |
| Month 6 | 77 | 85 | 0 | 0 | 0.459 | 0.4288 | 0.5493 |
| Month 9 | 90 | 90 | 0 | 0 | 0.3729 | 0.6412 | 0.7539 |

#### Effects on depression (as measured via the PHQ2 scale)

Depressive symptoms were assessed via the 2-point Patient Health Questionnaire (PHQ-2) [[4]](https://paperpile.com/c/PiWPX3/CyERh) (see items 17-18 in the baseline HQ in Box 9). Wilcoxon and Mann-Whitney one-sided tests were conducted as above. No statistically significant effects were observed for either group at any milestone, and no significant difference was observed between the two groups. Table 5.6 summarises the results of this analysis.

**Table 5.6:** Analysis of depressive symptoms. Reported p-values are for one-sided hypotheses (corresponding to a decrease in severity of depressive symptoms).

| **Milestone** | **No. Observations (Control)** | **No. Observations (Intervention)** | **Median score difference (Control)** | **Median score difference (Intervention)** | **Wilcoxon p-value (Control)** | **Wilcoxon p-value (Intervention)** | **Mann-Whitney p-value** |
| --- | --- | --- | --- | --- | --- | --- | --- |
| Month 3 | 96 | 99 | 0 | 0 | 0.576 | 0.077 | 0.1341 |
| Month 6 | 77 | 85 | 0 | 0 | 0.3484 | 0.498 | 0.7167 |
| Month 9 | 90 | 90 | 0 | 0 | 0.1869 | 0.296 | 0.4795 |

#### Effects on work-related outcomes

Several work-related outcomes were investigated: work efficiency (see item 24 in the Baseline HQ in Box 9), work quality (item 23), perceived workload (item 22), work satisfaction (item 25), and work autonomy (item 26). Wilcoxon and Mann-Whitney tests were applied as above. Table 5.7 summarises the results of these analyses. In general, there was little evidence of effects on work outcomes. Specifically, no significant within-group or between-group differences were observed for work quality, perceived increase in workload, work satisfaction, or work autonomy. For work quality, the same was true, except for a significant effect reported in the intervention group at month 9, with no corresponding between-group difference at that milestone. Perceived workload decrease also similarly demonstrated a significant effect in the intervention group for milestone 3, but again without a corresponding group difference at the same milestone. Table 5.7 summarises the results of these analyses.

Table 5.7: Analysis of work-related outcomes. Reported p-values are for one-sided outcomes.

| **Milestone** | **No. Observations (Control)** | **No. Observations (Intervention)** | **Median score difference (Control)** | **Median score difference (Intervention)** | **Wilcoxon p-value (Control)** | **Wilcoxon p-value (Intervention)** | **Mann-Whitney p-value** |
| --- | --- | --- | --- | --- | --- | --- | --- |
| **Work efficiency** | | | | | | | |
| Month 3 | 96 | 99 | 0 | 0 | 0.6129 | 0.2816 | 0.2931 |
| Month 6 | 77 | 85 | 0 | 0 | 0.6724 | 0.5778 | 0.4061 |
| Month 9 | 90 | 90 | 0 | 0 | 0.0567 | 0.0094 | 0.2073 |
| **Work quality** | | | | | | | |
| Month 3 | 96 | 99 | 0 | 0 | 0.71 | 0.155 | 0.0711 |
| Month 6 | 77 | 85 | 0 | 0 | 0.444 | 0.8221 | 0.7928 |
| Month 9 | 90 | 90 | 0 | 0 | 0.5696 | 0.1472 | 0.1907 |
| **Perceived Workload (increase)** | | | | | | | |
| Month 3 | 96 | 99 | 0 | 0 | 0.5769 | 0.9926 | 0.9075 |
| Month 6 | 77 | 85 | 0 | 0 | 0.8063 | 0.8545 | 0.6003 |
| Month 9 | 90 | 90 | 0 | 0 | 0.739 | 0.8993 | 0.6179 |
| **Perceived Workload (decrease)** | | | | | | | |
| Month 3 | 96 | 99 | 0 | 0 | 0.4287 | 0.0076 | 0.093 |
| Month 6 | 77 | 85 | 0 | 0 | 0.1976 | 0.1497 | 0.4011 |
| Month 9 | 90 | 90 | 0 | 0 | 0.2647 | 0.1027 | 0.3833 |
| **Work satisfaction** | | | | | | | |
| Month 3 | 96 | 99 | 0 | 0 | 0.2385 | 0.7982 | 0.898 |
| Month 6 | 77 | 85 | 0 | 0 | 0.7821 | 0.975 | 0.8944 |
| Month 9 | 90 | 90 | 0 | 0 | 0.2387 | 0.9341 | 0.9308 |
| **Work autonomy** | | | | | | | |
| Month 3 | 96 | 99 | 0 | 0 | 0.1739 | 0.622 | 0.6845 |
| Month 6 | 77 | 85 | 0 | 0 | 0.6785 | 0.9709 | 0.713 |
| Month 9 | 90 | 90 | 0 | 0 | 0.1303 | 0.562 | 0.875 |

#### Collective results following Bonferroni-Holm correction

Significance of all results above was re-evaluated using the Bonferroni-Holm correction procedure, testing at a family-wise error rate of 0.05 across 36 tests (per test category). This established that no effects persisted as significant following the correction procedure.

### Box 6: Why Milestone Choice Matters

#### User-Level Case Studies

The individual user journeys below demonstrate why Sync Date is the only valid anchor for baseline measurement. In each case, the gap between system-side milestones (First App Open, Split Date) and actual behavioural engagement (Sync Date) would severely contaminate baseline measurements if ignored.

**Example 1**: Late Syncer with 3-Month Gap

This user opened the app (blue dashed line) in September 2024 but did not sync their tracker (green line) until December 2024 (a 3-month gap). If we used First App Open as T=0, these 3 months of backfilled data (pink dots) would be misclassified as "treatment period" when the user was not yet engaged. Note the clear behavioural shift: Mean Pre-Sync = 2,098 steps vs. Mean Post-Sync = 6,679 steps (+219% increase), confirming that Sync Date marks the true behavioural activation.

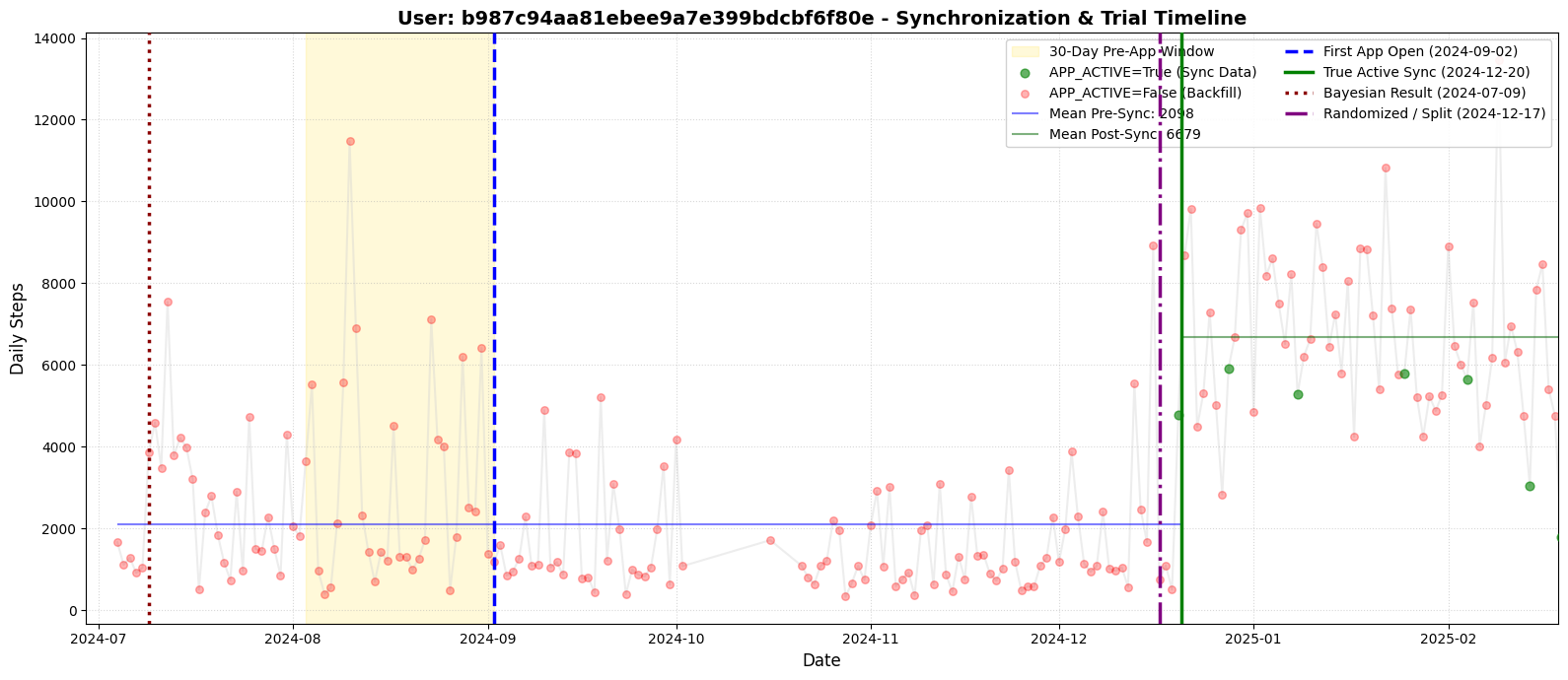

**Example 2**: Variable Tracking with Clear Sync Anchor

The Sync Date (green line) marks the start of consistent tracking. The gap between Sync and Split dates shows why system-side milestones misrepresent the user's actual behavioural start.

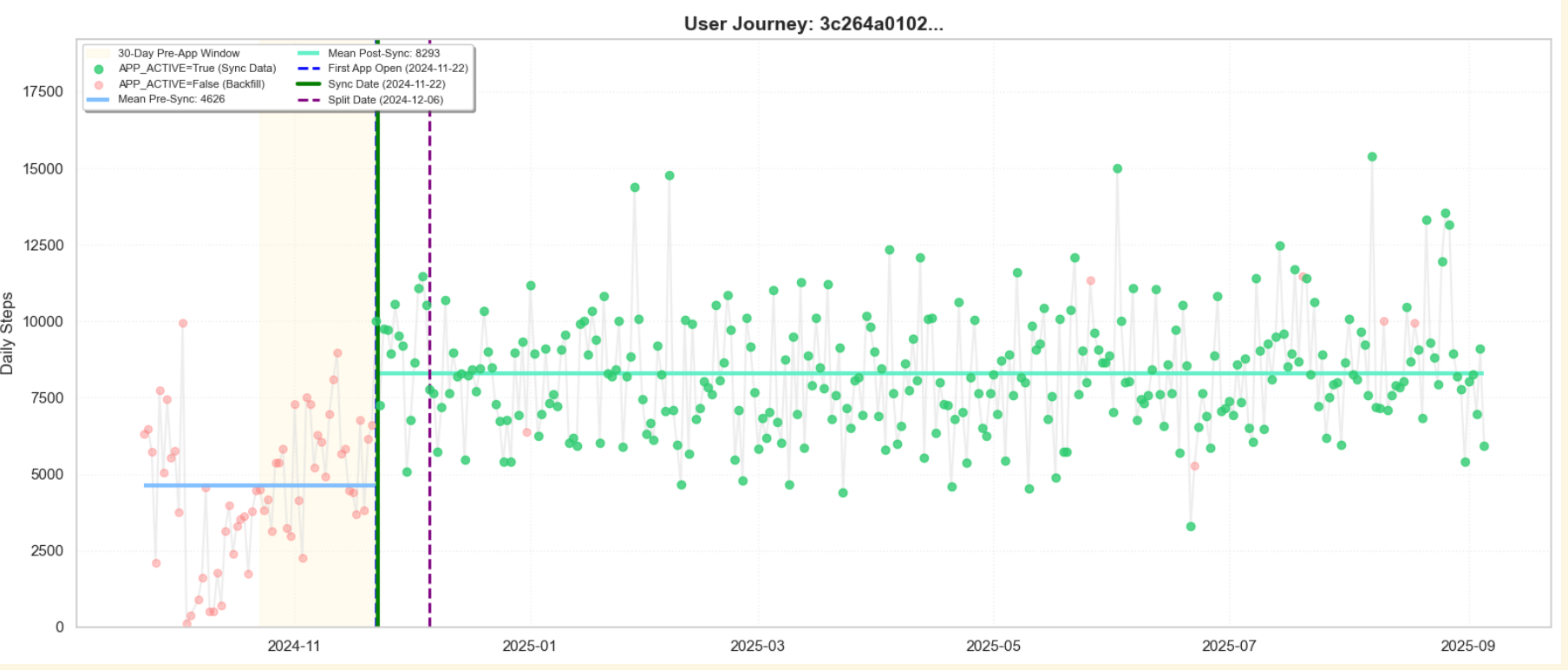

**Example 3:** Late Syncer — Why First App Open Fails as T=0

This user opened the app (blue dashed line) in December 2024 but did not sync their tracker (green line) until February 2025 (a 2-month gap). If we used First App Open or Split Date as T=0, this user would be classified as having no valid baseline as there is no tracking data prior to those dates. However, by adopting Sync Date as our temporal anchor, the backfilled data prior to sync becomes a valid baseline measurement. This is methodologically sound because the user was not yet exposed to the intervention: they could not see their steps, earn rewards, or engage with gamification features until the moment of sync. The backfilled historical data, therefore, represents their genuine pre-intervention activity level.

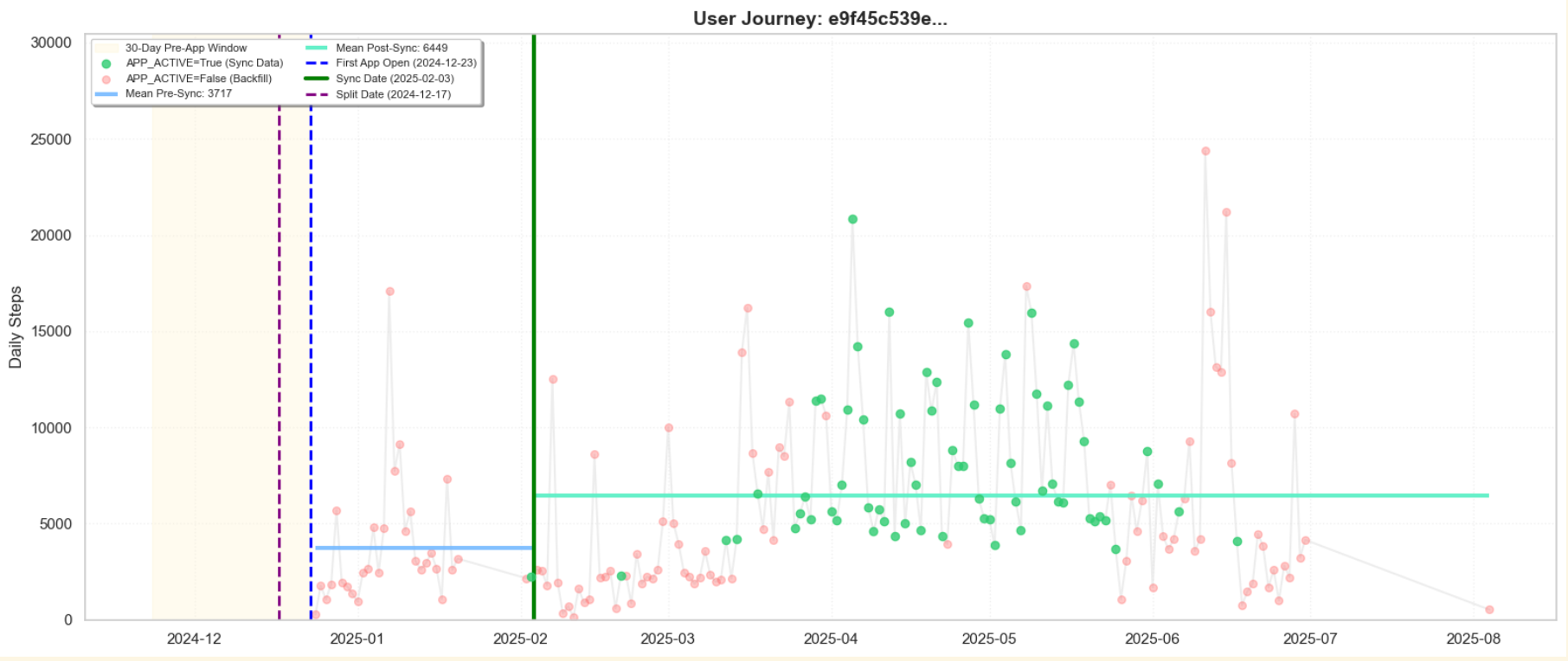

#### Novelty effect spike analysis

The main manuscript presents a sensitivity analysis, where participants are classified according to the quality of their ‘sync’ and surrounding activity, into ‘High Quality Sync’, ‘Fragmented’, and ‘De Novo Tracker’ user cohorts. Additionally, 3 temporal anchors were considered for defining baseline evaluation periods: ‘First App Open’, ‘Split Date’, and ‘Sync Date’. Table 6.1 explores how baseline steps change for each combination of cohort and temporal anchor, as well as when step counts peak and on what day within the baseline period. As explained in the main manuscript, the findings suggest a potential novelty effect following app exposure and justify using the Sync Date as a temporal anchor as a sensible alternative baseline definition, with potentially more meaningful results than the per-protocol definition proposed in the original study’s pre-registration protocol.

**Table 6.1:** Novelty effect spike analysis by cohort and anchor

| **Cohort** | **Anchor** | **Baseline** | **Peak** | **Peak Day** | **Spike** |
| --- | --- | --- | --- | --- | --- |
| High-Quality Sync | Sync Date | 7642 | 8486 | 4 | +844 |
| High-Quality Sync | Split Date | 7948 | 8451 | 11 | +502 |
| High-Quality Sync | App Open Date | 7664 | 8572 | 5 | +908 |
| Fragmented | Sync Date | 7407 | 9038 | 10 | +1631 |
| Fragmented | Split Date | 7569 | 8842 | 3 | +1273 |
| Fragmented | App Open Date | 7436 | 8800 | 11 | +1364 |
| De Novo Tracker | Sync Date | 2993 | 7194 | 14 | +4202 |
| De Novo Tracker | Split Date | 6158 | 7348 | 4 | +1190 |
| De Novo Tracker | App Open Date | 4330 | 6793 | 5 | +2463 |

### Box 7: Estimating Health Risk Reduction from steps via UK Biobank

#### Characteristics of the trained Cox Proportional Hazards model

A Cox Proportional Hazards model predicting risk of all-cause mortality was fitted on UK Biobank data. The predictors used for this model were Age, Sex, average daily step-count over the period of one week, subjective average daily sleep duration over 4 weeks, and BMI. The Table below shows the results of fitting the Cox model to the UK Biobank dataset with respect to the above predictors.

**Table 7.1:** Cox Proportional Hazards model results after fitting on the UK Biobank dataset, showing only statistically significant predictors.

|  | **coef** | **SE** | **coef lower 95%** | **coef upper 95%** | **p-value** |
| --- | --- | --- | --- | --- | --- |
| **Age (continuous variable)** | 0.110 | 0.0027 | 0.104 | 0.115 | < 0.001 |
| **Sex: ‘Female’ (reference: ‘Male’)** | -0.537 | 0.0343 | -0.604 | -0.470 | < 0.001 |
| **Steps: ‘5,000-8,000’ (reference: ‘<5,000’)** | -0.498 | 0.0962 | -0.687 | -0.310 | < 0.001 |
| **Steps: ‘8,000-10,000’ (reference: ‘<5,000’)** | -0.588 | 0.1123 | -0.808 | -0.368 | < 0.001 |
| **Steps: ‘10,000-12,000’ (reference: ‘<5,000’)** | -0.707 | 0.1404 | -0.982 | -0.432 | < 0.001 |
| **Steps: ‘>12,000’ (reference: ‘<5,000’)** | -0.834 | 0.1587 | -1.145 | -0.523 | < 0.001 |
| **Sleep: ‘Short Sleep (≤6h)’ (reference: 'Recommended Sleep (6-9h)')** | 0.154 | 0.0678 | 0.022 | 0.287 | 0.023 |
| **BMI: ‘Underweight’ (reference: ‘Normal’)** | 0.614 | 0.1917 | 0.238 | 0.989 | 0.001 |
| **BMI: ‘Obese’ (reference: ‘Normal’)** | 0.408 | 0.0462 | 0.317 | 0.498 | < 0.001 |

It is clear from the trained model coefficients that average daily step counts above 5000 are generally associated with lower mortality risk. Additionally, reduced sleep and deviations from normal BMI were associated with increased mortality risk, consistent with standard academic literature on the topic [[5]](https://paperpile.com/c/PiWPX3/xnnR).

#### Mapping participant steps to partial hazard using the trained Cox model

Table 7.2 shows the results of fitting an LMM model to partial hazards generated by plugging participant data (e.g., steps, age, gender) into the Cox model.

**Table 7.2.** LMM results for predicted partial hazard.

| **Effect** | **Coef.** | **Std.Err.** | **P>\|z\|** | **One-sided p-value** |
| --- | --- | --- | --- | --- |
| Intercept | −0.952 | 0.044 | < 0.001 |  |
| Timepoint: 'Month 3' (reference: 'base') | −0.009 | 0.006 | 0.121 |  |
| Timepoint: 'Month 5' (reference: 'base') | −0.002 | 0.006 | 0.687 |  |
| Timepoint: 'Month 7' (reference: 'base') | 0.002 | 0.006 | 0.718 |  |
| Timepoint: 'Month 9' (reference: 'base') | 0.007 | 0.006 | 0.261 |  |
| Group: 'Intervention' (reference: 'Control') | 0.001 | 0.024 | 0.954 |  |
| Sex: 'Male' (reference: 'Female') | 0.137 | 0.023 | < 0.001 |  |
| Month 3 × Intervention | −0.007 | 0.008 | 0.394 | 0.197 |
| Month 5 × Intervention | −0.018 | 0.008 | 0.032 | 0.016 |
| Month 7 × Intervention | −0.009 | 0.009 | 0.287 | 0.144 |
| Month 9 × Intervention | −0.026 | 0.009 | 0.003 | 0.002 |
| Age (continuous) | 0.029 | 0.001 | < 0.001 |  |
| Group Var (random intercept variance) | 0.096 | 0.078 | — |  |

### Box 8: LMM results – Alternative baseline definition analyses.

**Table 8.1:** Baseline characteristics of participants with a valid alternative baseline

| **Variable** | **Intervention (n=407)** | **Control (n=432)** | **Total (n=839)** | **p-value*** |
| --- | --- | --- | --- | --- |
| Age (mean, SD) | 39.6 (11.2) | 39.5 (11.5) | 39.5 (11.3) | 0.931 |
| Gender (% female) | 204 (50%) | 219 (51%) | 423 (50%) | 0.869 |
| Average steps (mean, SD) | 7473.6 (3345) | 7723.3 (3795) | 7602.1 (3586) | 0.314 |

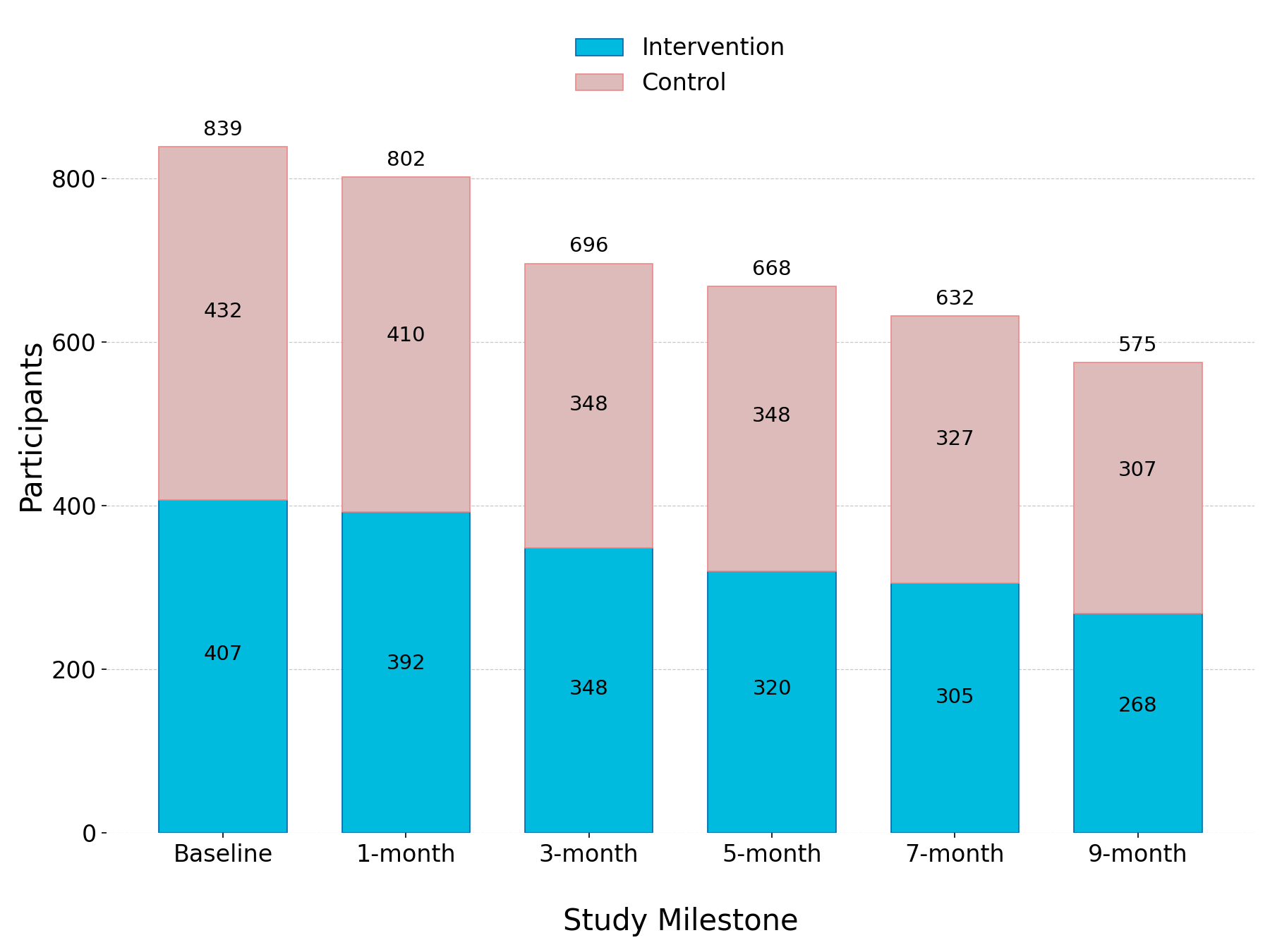

**Figure 8.1:** Participant retention using the alternative baseline definition

| **A** | **B** |
| --- | --- |
| 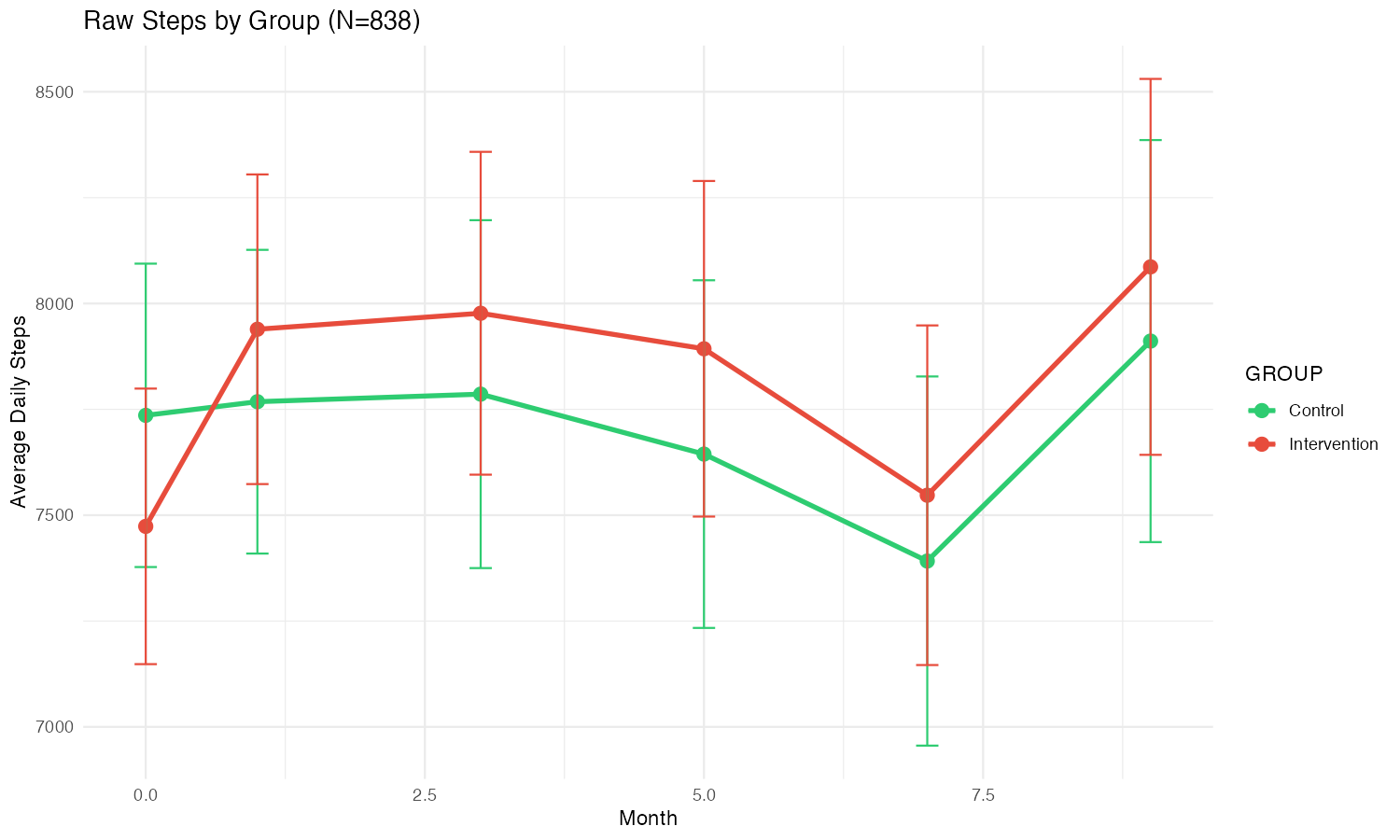 | 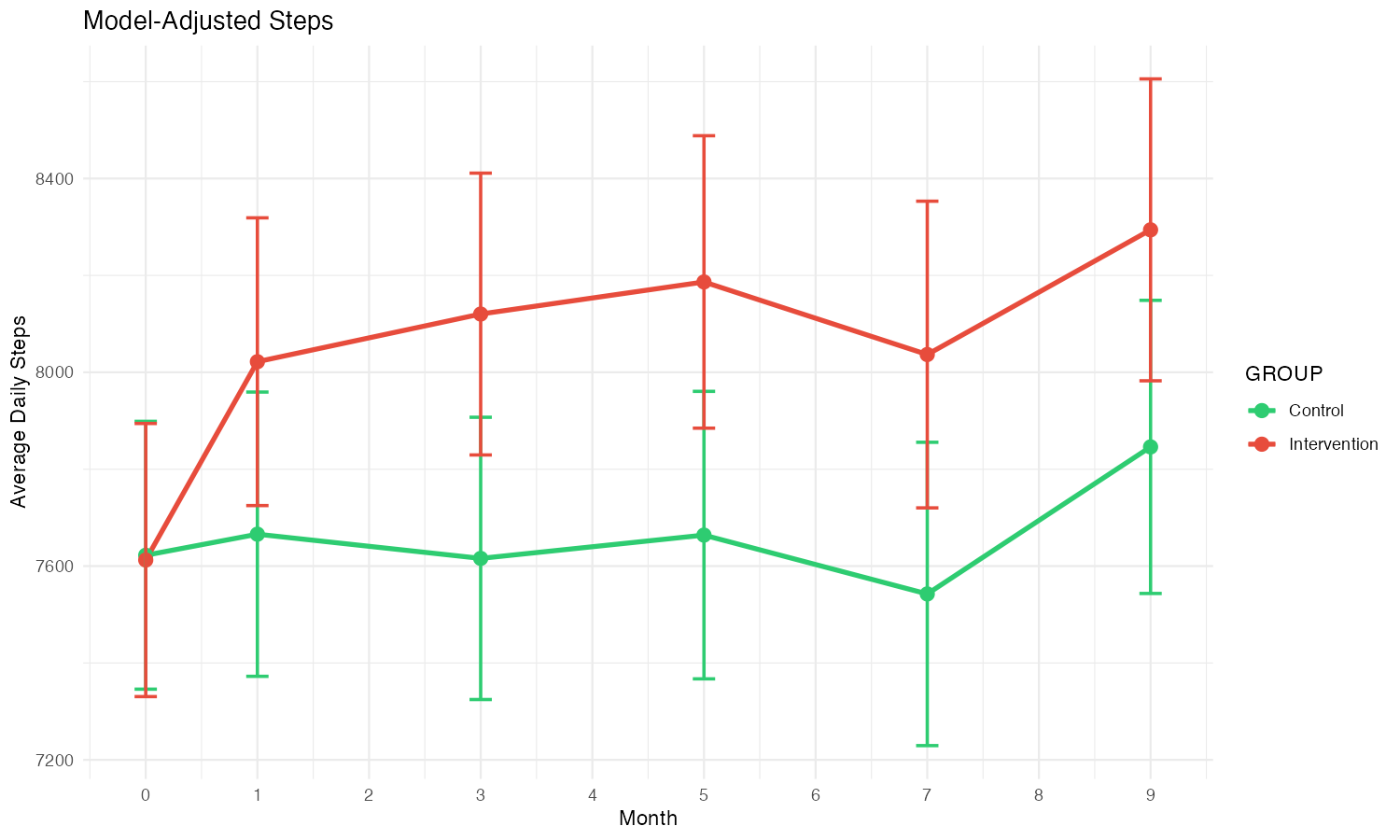 |

**Figure 8.2:** (A) Unadjusted and (B) model-adjusted (via Estimated Marginal Means) mean daily step counts by time × group (±1 SE), holding season and baseline at reference values, when using the alternative baseline definition.

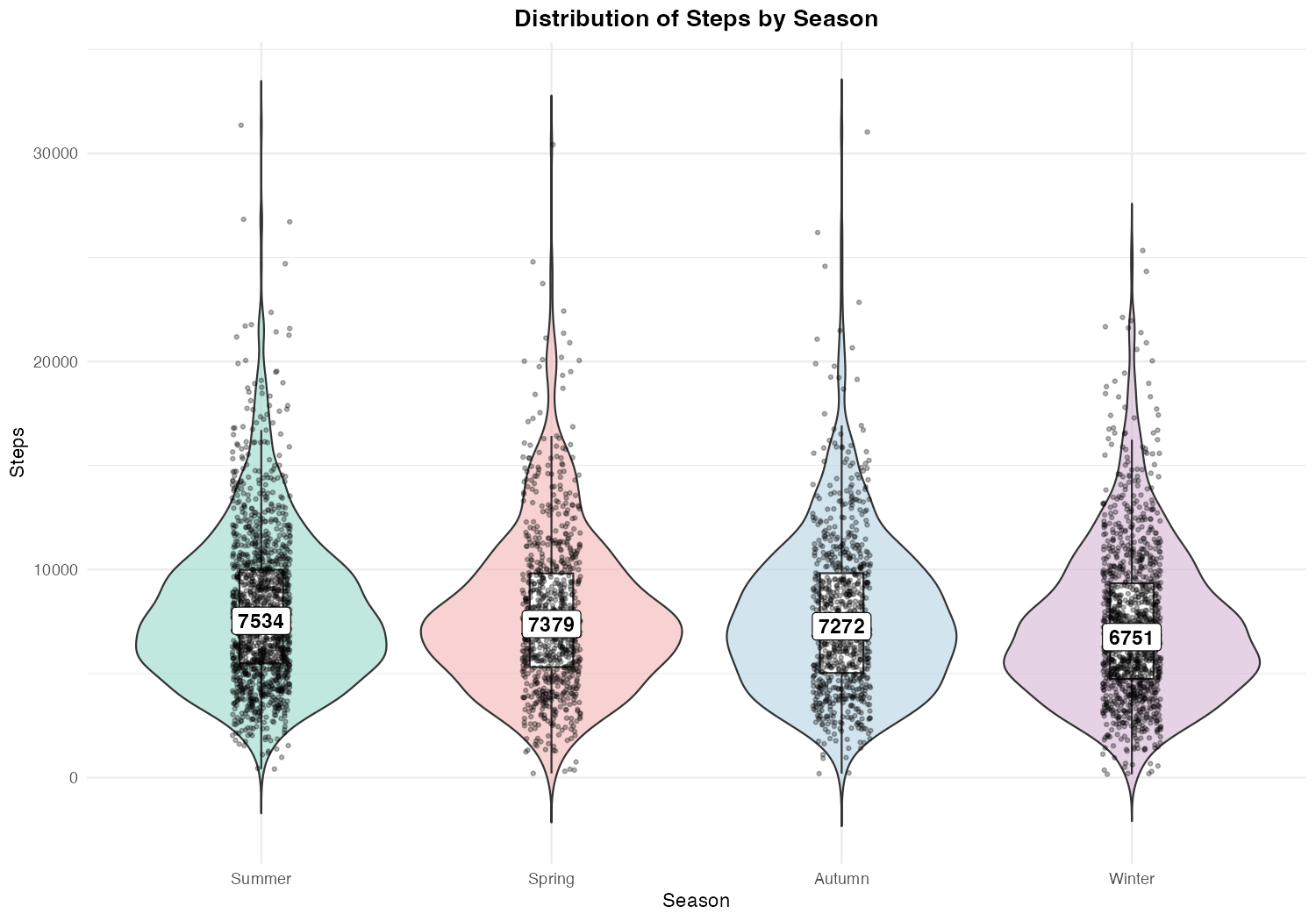

**Figure 8.3:** Distribution of steps by season under the alternative baseline definition

**Table 8.2:** LMM results for mean daily step count using alternative baseline definition.

| **Term** | **Estimate** | **Std.Err.** | **P>\|z\|** | **One-sided p-value** |
| --- | --- | --- | --- | --- |
| (Intercept) | 6694.24 | 1,498.34 | <0.001 |  |
| Milestone: Month 1 (reference: baseline) | 44.135 | 137.748 | 0.749 |  |
| Milestone: Month 3 | -6.322 | 146.752 | 0.966 |  |
| Milestone: Month 5 | 40.688 | 156.148 | 0.794 |  |
| Milestone: Month 7 | -81.487 | 157.517 | 0.605 |  |
| Milestone: Month 9 | 220.965 | 151.259 | 0.144 |  |
| Group: Intervention (reference: Control) | -13.219 | 165.761 | 0.936 |  |
| Baseline (centred) | 0.8183 | 0.017 | <0.001 |  |
| Season: Spring (reference: Summer) | -373.322 | 104.054 | <0.001 |  |
| Season: Autumn | -616.115 | 114.137 | <0.001 |  |
| Season: Winter | -939.436 | 103.384 | <0.001 |  |
| BMI | -28.728 | 12.112 | 0.018 |  |
| Month 1 × Intervention | 366.182 | 189.894 | 0.054 | 0.027 |
| Month 3 × Intervention | 514.957 | 197.258 | 0.009 | 0.005 |
| Month 5 × Intervention | 533.732 | 198.76 | 0.007 | 0.004 |
| Month 7 × Intervention | 505.415 | 202.7 | 0.013 | 0.007 |
| Month 9 × Intervention | 459.085 | 208.996 | 0.028 | 0.014 |

### Box 9 - Questionnaires used in the study

#### Baseline Health Questionnaire

**Baseline Health, Lifestyle and Productivity Assessment for the University of Essex & YuLife Gamification Intervention Study**

This questionnaire is designed to gather information about your health, lifestyle, and productivity habits. Your responses will help us understand your current state and track any changes over the course of the study.

The questionnaire includes questions about your physical activity, dietary habits, mental health, and work productivity. It should take about 5 - 10 minutes to complete. All your responses will be kept confidential, anonymised for analysis and will not be shared with your employer.

Thank you for your time and contribution to this important research.

If you have any questions, concerns, or need more information regarding any aspect of the study, please feel free to contact the Study Lead, Dr Abbas Salami, at.

*1. What is your gender?*

- Male
- Female
- Non-binary
- Other
- Prefer not to say

*2. Do you currently use a wearable fitness tracker or smartwatch, or are you interested in using one to monitor your health?*

- Yes, I currently use one
- No, but I am interested in using one
- No, and I am not interested in using one

*3. What is your ethnicity?*

- Asian/Asian British
- Black/African/Caribbean/Black British
- Mixed/Multiple ethnic groups
- Other ethnic group
- White
- Prefer not to say

*4. Please enter your height in your preferred unit and leave the other box empty.*

- Metric (centimeters)
- Imperial (feet & inches)

*5. Please enter your weight in your preferred unit and leave the other box empty.*

- Metric (kilograms)
- Imperial (stones & pounds)

*6. Do you smoke or consume tobacco or nicotine in any form?*

- Yes, cigarettes or similar tobacco products
- Yes, I use vaping devices with nicotine
- Yes, nicotine patches/gum (i.e. not from smoking or vaping)
- Yes, other
- No

*7. How frequently do you consume alcohol?*

- Never
- Special occasions only
- 1-3 times a month
- 1-2 times a week
- 3-4 times a week
- Daily or almost daily

*8. Do you have any injuries, musculoskeletal conditions, other health issues, or something else that could limit your physical activity?*

- Yes
- No

*9. Do you have any dietary restrictions or preferences? (can select multiple)*

- Vegetarian
- Vegan
- Gluten-free
- Have certain food allergies
- Religious/Cultural restrictions
- Other
- No restrictions

*10. Do you currently practise meditation or mindfulness exercises?*

- Regularly (Multiple times a week)
- Occasionally (2-4 times a month)
- Rarely (Monthly or less)
- Never

*11. Over the past month, how many days in a typical week do you engage in 30 minutes or more of moderate intensity physical activity or exercise? (Examples of moderate intensity activities include going to the gym, running, playing sports like football, tennis, cycling, swimming, etc.)*

- 0 days
- 1-2 days
- 3-4 days
- 5-7 days

*12. Are you currently taking any medications regularly?*

- Yes
- No

*13. In the last month, how often have you felt that you were unable to control the important things in your life?*

- Never
- Almost Never
- Sometimes
- Fairly Often
- Very Often

*14. In the last month, how often have you felt confident about your ability to handle your personal problems?*

- Never
- Almost Never
- Sometimes
- Fairly Often
- Very Often

*15. In the last month, how often have you felt that things were going your way?*

- Never
- Almost Never
- Sometimes
- Fairly Often
- Very Often

*16. In the last month, how often have you felt difficulties were piling up so high that you could not overcome them?*

- Never
- Almost Never
- Sometimes
- Fairly Often
- Very Often

*17. Over the last 2 weeks, how often have you felt little interest or pleasure in doing things?*

- Not at all
- Several days
- More than half the days
- Nearly every day

*18. Over the last 2 weeks, how often have you felt down, depressed or hopeless?*

- Not at all
- Several days
- More than half the days
- Nearly every day

*19. Over the last 2 weeks, how often have you felt nervous, anxious or on edge?*

- Not at all
- Several days
- More than half the days
- Nearly every day

*20. Over the last 2 weeks, how often have you not been able to stop or control worrying?*

- Not at all
- Several days
- More than half the days
- Nearly every day

*21. In the last 2 weeks, how many hours did you sleep over a typical 24 hour period?*

*22. Over the past month, how rewarding did you find your work?*

- Not rewarding at all
- Somewhat rewarding
- Neutral
- Quite rewarding
- Extremely rewarding

*23. How much control did you feel you had over your job in the past month?*

- No control at all
- Little control
- Neutral
- Considerable control
- Total control

*24. How would you describe your overall efficiency at work over the past month?*

- Very inefficient
- Somewhat inefficient
- Neutral
- Quite efficient
- Very efficient

*25. How would you rate the overall quality of your work over the past month?*

- Very poor
- Somewhat poor
- Neutral
- Quite good
- Very good

26. How would you describe the overall amount of work you did in the past month?

- Very little
- Somewhat little
- Neutral
- Quite a lot
- A lot

#### Follow-up questionnaires

**Quarterly Health, Lifestyle and Productivity Assessment for the University of Essex & YuLife Gamification Intervention Study**

This questionnaire is designed to gather information about your health, lifestyle, and productivity habits. Your responses will help us understand your current state and track any changes over the course of the study.

The questionnaire includes questions about your physical activity, dietary habits, mental health, and work productivity. It should take about 5 - 10 minutes to complete. All your responses will be kept confidential, anonymised for analysis and will not be shared with your employer.

Thank you for your time and contribution to this important research.

If you have any questions, concerns, or need more information regarding any aspect of the study, please feel free to contact the Study Lead, Dr Abbas Salami, at.

*1. Please enter your weight in your preferred unit and leave the other box empty.*

- Metric (kilograms)
- Imperial (stones & pounds)

*2. Do you smoke, or have you ever smoked cigarettes?*

- Never
- Former smoker (less than 20 cigarettes/day)
- Former smoker (20 or more cigarettes/day)
- Current smoker (less than 20 cigarettes/day)
- Current smoker (20 or more cigarettes/day)

*3. Have you smoked or consumed tobacco or nicotine in any form over the past month?*

- Yes, cigarettes or similar tobacco products
- Yes, I use vaping devices with nicotine
- Yes, nicotine patches/gum (i.e. not from smoking or vaping)
- Yes, other
- No

*4. How frequently have you consumed alcohol over the past 3 months?*

- Never
- Special occasions only
- 1-3 times a month
- 1-2 times a week
- 3-4 times a week
- Daily or almost daily

*5. Did you have any injuries, musculoskeletal conditions, other health issues, or something else that could have limited your physical activity over the past 3 months?*

- Yes
- No

*6. Do you have any dietary restrictions or preferences? (can select multiple)*

- Vegetarian
- Vegan
- Gluten-free
- Have certain food allergies
- Religious/Cultural restrictions
- Other
- No restrictions

*7. Over the past month, how many days in a typical week do you engage in 30 minutes or more of moderate intensity physical activity or exercise? (Examples of moderate intensity activities include going to the gym, running, playing sports like football, tennis, cycling, swimming, etc.)*

- 0 days
- 1-2 days
- 3-4 days
- 5-7 days

*8. Are you currently taking any medications regularly?*

- Yes
- No

*9. In the last month, how often have you felt that you were unable to control the important things in your life?*

- Never
- Almost Never
- Sometimes
- Fairly Often
- Very Often

*10. In the last month, how often have you felt confident about your ability to handle your personal problems?*

- Never
- Almost Never
- Sometimes
- Fairly Often
- Very Often

*11. In the last month, how often have you felt that things were going your way?*

- Never
- Almost Never
- Sometimes
- Fairly Often
- Very Often

*12. In the last month, how often have you felt difficulties were piling up so high that you could not overcome them?*

- Never
- Almost Never
- Sometimes
- Fairly Often
- Very Often

*13. Over the last 2 weeks, how often have you felt little interest or pleasure in doing things?*

- Not at all
- Several days
- More than half the days
- Nearly every day

*14. Over the last 2 weeks, how often have you felt down, depressed or hopeless?*

- Not at all
- Several days
- More than half the days
- Nearly every day

*15. Over the last 2 weeks, how often have you felt nervous, anxious or on edge?*

- Not at all
- Several days
- More than half the days
- Nearly every day

*16. Over the last 2 weeks, how often have you not been able to stop or control worrying?*

- Not at all
- Several days
- More than half the days
- Nearly every day

*17. In the last 2 weeks, how many hours did you sleep over a typical 24 hour period?*

*18. Over the past month, how rewarding did you find your work?*

- Not rewarding at all
- Somewhat rewarding
- Neutral
- Quite rewarding
- Extremely rewarding

*19. How much control did you feel you had over your job in the past month?*

- No control at all
- Little control
- Neutral
- Considerable control
- Total control

*20. How would you describe your overall efficiency at work over the past month?*

- Very inefficient
- Somewhat inefficient
- Neutral
- Quite efficient
- Very efficient

*21. How would you rate the overall quality of your work over the past month?*

- Very poor
- Somewhat poor
- Neutral
- Quite good
- Very good

*22. How would you describe the overall amount of work you did in the past month?*

- Very little
- Somewhat little
- Neutral
- Quite a lot
- A lot

#### Dynamic Health Questionnaire

The Dynamic Health Questionnaire (DHQ) used in the study consisted of around 500 questions, divided into ‘topics’. Users were shown only a handful of questions at pre-specified intervals throughout the study (see the main text for details). The table below shows the topics covered in the DHQ, along with a small, illustrative sample of the kinds of questions that would be asked for each topic.

*Sleep:*

- How many hours of sleep did you get last night?
- Are you a morning or night person?
- Within the last two weeks, have you been made aware that you snore? If so, how severe is it?

*Stress management:*

- In the past 6 months, how often have you felt stressed?
- In the past month, have there been recurring stress triggers in your life?
- In the past week, have you noticed any patterns in your physical environment that may trigger stress, such as noise, clutter, or lighting, and what changes have you made to reduce their impact?

*Attention and Focus:*

- In the past week, how often did you manage to stay focused on important tasks without getting sidetracked?
- What are some methods you use to boost your focus when working on something that needs all your attention?
- Do you stick to your focus methods?

*Social connections:*

- How often do you socialise with friends, family, or community groups?
- In the past week, have you had any meaningful conversations with someone close to you?
- In the past week, how satisfied have you been with the quality of your social interactions?

*Tobacco use:*

- Have you used nicotine products more than once in the past 6 months?
- Which of the following did you use yesterday? (Select all that apply)
- Are you currently seeking to quit nicotine use?

*Alcohol Consumption:*

- Have you had an alcoholic drink in the past 6 months?
- Are you interested in receiving some advice about alcohol consumption? Just checking in to see if you're open to some helpful tips!
- What is your most preferred alcoholic drink?

*Diet:*

- Are you happy with your current diet?
- In the last month, did you regularly eat fast food or processed foods?
- In the last month, how often did you eat healthy and balanced meals?

*Limiting caffeine and sugar intake:*

- Did you monitor how much caffeine you drank yesterday?
- Rather than caffeinated or sugary drinks, did you pick alternative drink choices yesterday such as herbal tea or water?
- If you like sugary treats, did you take any steps yesterday to cut back on them?

*Mobile Phone & Social Media Use:*

- How do you feel after spending a significant amount of time on your mobile phone or browsing social media?
- In the past month, have you often found yourself reaching for your phone without a specific purpose or out of habit?
- In the past 2 months, you felt the need to take a break from your mobile phone or social media for mental health reasons?

*Hydration:*

- During the day, do you keep track of how many non-alcoholic drinks you drink?
- If you drank alcohol yesterday, did you also drink water with it?
- It is recommended you drink 8-10 glasses of water (2-2.5 litres) a day. Yesterday, did you drink that amount of water?

*Social Awareness:*

- In the past month, have you been told that something you said or did might have made others feel uncomfortable in social settings?
- Do you feel comfortable addressing behaviours of others, such as coworkers, friends or family, when you find them inappropriate or discomforting?
- Looking back over the past month, do you think you've made changes to your behavior in social settings based on feedback you've received?

*Behavioral Impact:*

- After a setback or receiving feedback, do you pause to reflect on your actions, thinking about how you could improve future interactions or results?
- In the past month, have your behaviour or actions caused any personal or professional setbacks?
- When encountering conflicts, how do you usually resolve them?

*Cultivating resilience:*

- In the past month, when you faced challenges or setbacks, did you try to be understanding and kind towards yourself, rather than overly self-critical?
- Did you do any activities yesterday to help manage stress?
- Do you find yourself finding silver linings when faced with negative thoughts or experiences?

*Depression:*

- In the past month, have you experienced any periods of feeling depressed or down?
- Do you still participate in activities you enjoy doing?
- Do you have any personal methods that help you cope with feeling down or low on a daily basis?

*Physical activity:*

- Do you engage in any team sports or group physical activities?
- Do you do physical activities that improve your strength, such as weight lifting, resistance band exercises, or bodyweight exercises?
- Do you do any physical activities that improve balance and coordination, such as yoga, tai chi, or dance?

*Building self-awareness and introspection:*

- In the past month, did you actively seek feedback from your peers or people you trust?
- If you ask people for feedback, what do you usually look for in their answers?
- Do you set personal goals around health and wellbeing, and track your progress or adjust them if you need to?

*Pain and Musculoskeletal (MSK) conditions:*

- Can you usually recognize signs of overwork in your body?
- Do you adjust your plans when you find that your body is overworked?
- Have you ever been diagnosed with rheumatoid arthritis?

*Mental health care:*

- Yesterday, did you set aside time for self-care activities to manage your stress or boost your wellbeing?
- Do you have access to professional help for mental health concerns if needed?
- If you've ever struggled with mental health concerns, have you sought professional help?

*Social Norms:*

- Yesterday, were there any situations where your opinion of what was socially appropriate was different from the people around you?
- Does your opinion on what is socially appropriate often seem to differ from the opinions of people around you?
- Over the past month, have you tried to change your behaviour to match the norms of the people around you?

*Social Conduct:*

- In the past week, were there any moments when your behaviour might have been considered tactless or embarrassing? What have you done about them?
- How frequently do you find yourself in situations where your behaviour may have been considered to be tactless or embarrassing?
- If you were informed, or realised, that your behaviour was tactless or embarrassing, what would you do?

*Mindful eating:*

- Did you eat your meals and snacks yesterday without distractions, like using your phones, TV, or computer?
- Did you explore your food choices yesterday with a curious, open-minded attitude, focusing on appreciation rather than seeing it as a reward, punishment, or something to feel guilty about?
- Did you think about setting a positive goal for your eating yesterday? Maybe something like focusing on nourishing your body with yummy, wholesome foods and enjoying every bite without feeling like you're missing out or limiting yourself?

*Addressing unresolved emotional issues:*

- In the past week, did you think about your emotions? Were you aware of any unresolved issues or conflicts that might be affecting you?
- In the past week, did you seek support from trusted people in your life to discuss your feelings or for support on any unresolved emotional issues?
- If you have any unresolved emotional issues, how are you addressing them?
